# Renal Resistance During Hypothermic Machine Perfusion: A Scoping Review of Variability and Determinants, with a Meta-Analysis of Predictive Value for Transplant Outcomes

**DOI:** 10.1101/2025.04.18.25326058

**Authors:** Liliana Fonseca Buitrago, Laurence Verstraeten, Steffen Fieuws, Ina Jochmans

**Affiliations:** Lab of Abdominal Transplantation, Transplantation Research Group, Department of Microbiology, Immunology and Transplantation, KU Leuven, Leuven, Belgium; Interuniversity Institute for Biostatistics and Statistical Bioinformatics, Department of Public Health, KU Leuven, Leuven, Belgium; Abdominal Transplant Surgery, University Hospitals Leuven, Leuven, Belgium

## Abstract

**Background:** Renal resistance (RR) measured during hypothermic machine perfusion (HMP) is used to assess donor kidney quality and guide transplantation decisions. However, its clinical reliability and relationship with donor factors remain unclear.

**Methods:** This scoping review and meta-analysis evaluate the variability, determinants, and predictive value of RR during HMP. A systematic search of PubMed, Embase, Web of Science, and Cochrane Library (July 2024) identified 49 primary studies reporting RR in perfused human kidneys. The risk of bias was assessed using the ROBINS-I tool. Meta-analyses for the predictive value of RR were performed when ≥3 studies reported univariable associations for the same time point and outcome.

**Results:** Most studies had moderate to serious risk of bias. RR typically declined rapidly, stabilizing within 5 hours (range: 0.30–3.50 to 0.17–1.50 mmHg/mL/min), but patterns varied widely. Determinants included histology, donor characteristics, and perfusion additives, though evidence was inconsistent. A meta-analysis showed terminal RR was significantly associated with delayed graft function (odds ratio 2.49, 95% CI 1.49-4.18, I^2^=58%). While several studies proposed RR-thresholds, none were consistently validated, and heterogeneity in measurement timings and device settings limits comparability.

**Conclusion:** RR shows potential as a functional assessment parameter during HMP but is influenced by multiple technical and biological factors. Current evidence does not support the use of isolated RR-thresholds for organ acceptance. Standardized HMP protocols, trajectory modeling, and prospective studies are needed to clarify RR’s role in clinical decision-making.

**Funding:** This study was funded by a grant from the KU Leuven Research Council (C2M/23/051) and was preregistered at the Open Science Framework (DOI: 10.17605/OSF.IO/D8QYU).

## Introduction

As kidney transplant waiting lists continue to grow, the increasing reliance on higher-risk donor organs makes optimal organ preservation and selection more critical than ever. Hypothermic machine perfusion (HMP) has demonstrated several advantages over static cold storage, particularly in deceased donor kidney transplantation ^1–3^. These benefits include reduced risks of delayed graft function (DGF), potential improvements in graft survival, and enhanced cost-effectiveness.^1,4^

An additional advantage of HMP is the ability to evaluate the kidney in a dynamic environment before transplantation.^4^ One of the key parameters measured during HMP is renal resistance (RR), which reflects the impedance of the perfusate flow through the kidney’s vascular system. It is calculated by measuring flow at a given pressure while the kidney is connected to a perfusion circuit with pulsatile or continuous flow maintained by a pump. Numerous studies have explored the potential of RR as a predictor of post-transplant outcomes. Despite widespread interest, there is no consensus on what RR values signify, how they should be interpreted, or whether they should influence clinical decision-making – and a comprehensive synthesis of these studies is still lacking. Such a synthesis is essential to determine whether using RR as a criterion for discarding kidneys is clinically justified. Indeed, RR is currently used as a tool for decision-making in kidney transplantation.^5–9^ For example, in the United States, 33% of pumped kidneys were discarded in 2023, with kidneys exhibiting elevated RR (>0.30 mmHg/mL/min) having a 2 to 8-fold increased odds of discard.^10–12^

To address this gap, we conducted a scoping review and meta-analysis to: (1) characterize the typical trajectory and variability of RR during HMP, (2) examine the factors influencing RR, and (3) evaluate its predictive value for post-transplant outcomes.

## Methods

### Search strategy, study selection, and eligibility criteria

We searched PubMed, Embase, Web of Science, and Cochrane libraries from inception through July 15, 2024 (search strategies in Appendix S1). Studies were imported into Rayyan, where duplicates were removed using the “Detect Duplicates” tool.^13^ Additional records were identified by screening reference lists of included articles using Citation Chaser.^14^ To provide a comprehensive overview and identify knowledge gaps, we applied inclusive eligibility criteria (Appendix S2). Eligible articles reported perfusion parameters during HMP of human kidneys. At least two independent reviewers screened each record, resolving discrepancies through discussion. This scoping review was preregistered in the Open Science Framework.^15^

### Primary and Secondary objectives

The primary objective was to describe RR values and trajectories during HMP. Secondary objectives included identifying determinants of RR (i.e. factors that may influence RR) and evaluating associations between RR and post-transplant outcomes.

### Selection of primary study reports, data extraction, and effect measures

To avoid duplicate data, we identified one primary report per study based on sample size and publication date (Appendix S3). Related reports were included only if they reported additional outcomes not covered in the primary report. Data extraction followed a standardized protocol, capturing study characteristics, population and perfusion characteristics, and any reported correlations of RR with determinants and post-transplant outcomes like DGF, kidney function, graft survival (Fig. S6). For the latter, we extracted the odds ratio (OR), hazard ratio (HR), and the area under the receiver operating characteristic curve (AUC), from univariable and multivariable models. Renal vascular resistance data were also extracted. Point estimates were determined from available graphs using WebPlotDigitizer v.4.3 (Ankit Rohatgi, California, USA)^16^ unless the necessary data points were reported.^17^ For studies reporting RR in mmHg/mL/min/100 g, the values were converted to mmHg/mL/min using the mean kidney weight reported in the paper or an estimated average kidney weight of 141g (Appendix S4).

### Risk of bias and quality assessment

The ROBINS tool (Risk of Bias in Non-randomized Studies) was used to assess the bias risk and evaluate the quality of the included studies. The tool evaluated bias due to confounding, participant selection, intervention classification, deviations from intended interventions, missing data, outcome measurement, and reported results selection, providing a comprehensive evaluation of the studies included in this research.

### Data synthesis and Meta-analysis

Renal resistance values across studies were visualized as mean or median estimates at specific time points or the end of perfusion. Subgroup analyses were performed based on donor type and publication year, using 2000 as a cut-off point. This year corresponds to the introduction and widespread adoption of the LifePort perfusion device, which marked a shift toward more standardized machine perfusion practices. Studies proposing RR thresholds for clinical outcomes were further assessed for methodological robustness using predefined grading criteria (Appendix S6).

Meta-analyses were conducted only when at least three studies assessed RR at the same time point for the same outcome using univariable analysis (Appendix S5). Due to heterogeneity in measurement units, time points, and statistical methods, this criterion was only met for the association between terminal RR and DGF. A random-effects meta-analysis evaluated the association between terminal RR and DGF (using odds ratio’s [OR]) and its predictive accuracy (using AUC values). Study weights were assigned via the inverse-variance method and pooled estimates with their 95% confidence intervals (CIs) were calculated. Heterogeneity was assessed using the I^2^ statistic. To assess the robustness of the meta-analysis findings, a Leave-One-Out sensitivity analysis was performed by systematically excluding each study and recalculating the pooled OR and CI to determine if a single study disproportionally influenced the results. All analyses were conducted using Review Manager (RevMan, version 5.4; The Cochrane Collaboration, 2020).^18^

## Results

### Study Description and Quality Assessment

The systematic search identified 9187 articles (PubMed: 2956, Embase: 3195, Web of Science: 2777, and Cochrane Library: 259) of which 64 met the inclusion criteria. An additional 7 articles identified through references searching (Fig. S1). Of 71 articles with primary and secondary outcome data, 49 primary study reports were identified (Table S1).^2,5,8,12,19–85^ The full data extraction table can be accessed online.^86^

These 49 studies collectively reported on 17798 perfused kidneys over a nearly four-decade period (1988-2024). Most were conducted in the United States (20/49, 40%), followed by Eurotransplant countries (6/49, 12%), China (5/49, 10%), Italy (4/49, 8%), Japan and the United Kingdom (3/49, 6% each), Poland, Spain, and Canada (2/49, 4% each), and Brazil, France, and Romania (1/49, 2% each) (Table S2). Study designs were predominantly retrospective (30/49, 61%), with 33% using prospectively collected (16/49) and 6% comprising case series (3/49). Risk of bias was rated as critical (10/49, 20%), serious (21/49, 43%), or moderate (16/49 33%) in most studies, with only two studies (4%) judged to have low risk (Fig. 1, S5).

**Fig 1.**
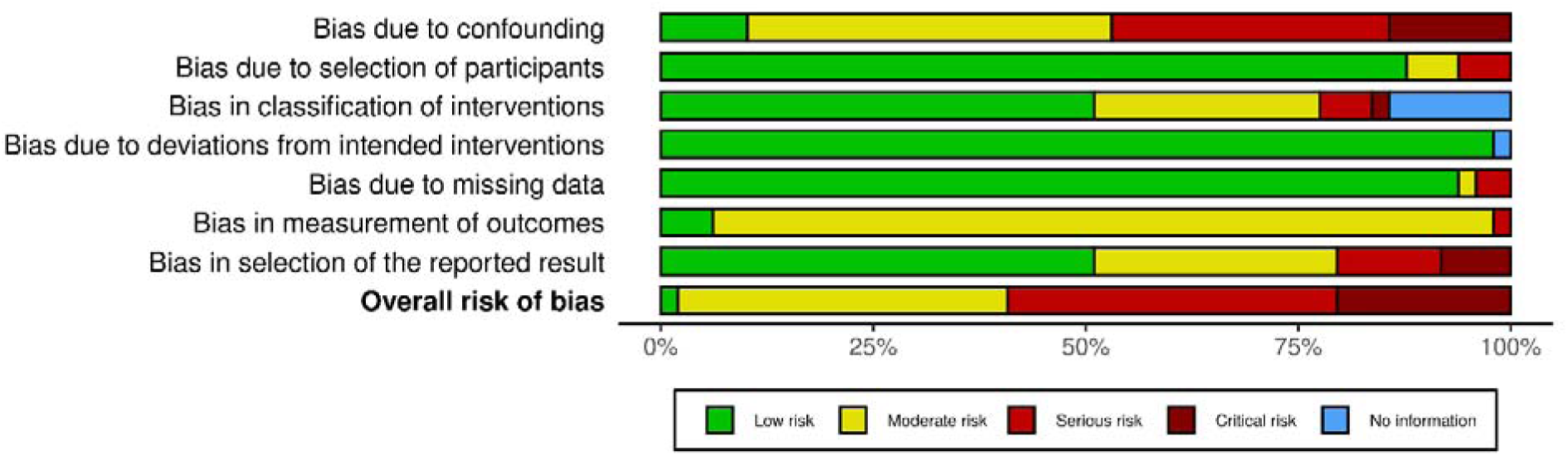
ROBINS I summary plot. This plot shows the risk of bias in non-randomized studies using the ROBINS-I tool. It evaluates bias across seven domains: confounding, participant selection, intervention classification, deviations from intended interventions, missing data, outcome measurement, and reported results. Each domain is color-coded to indicate the risk level: low, moderate, serious, or critical.

### Donor Characteristics and Preselection Criteria

The included studies encompassed a range of donor types. Kidneys were donated after brain death (DBD, (15/49, 30.6%), after circulatory death (DCD, 15/49, 30.6%), or both (13/49, 26.5%), while 10% of studies did not specify donor type (5/49). One study (2%) reported exclusively on living donors. Among DBD donors, 51% were standard criteria, 33% expanded criteria, and 16% were unspecified.

Seven studies applied preselection criteria based on perfusion parameters, using thresholds for flow, RR or RI (Table S3).^27,45,56,61,63,64,81^ For example, Mendez et al. discarded kidneys with a terminal RR >0.30 mmHg/mL/min,^27^ while Matillon et al. applied a 6-hour threshold >0.50 mmHg/mL/min.^45^

### Perfusion techniques and Device Variability

All kidneys were perfused under hypothermic conditions, typically 1-8°C, though some studies reported temperatures up to 12°C.^29^ Perfusion and donor warm ischemia times varied widely (Table S2). Nine different perfusion devices were used, of which only some are commercially available today (Table S4). LifePort was the most common (25/49, 51%), followed by RM3 (12%), MOX100 (10%), Gambro PF-3B (6%), and LPS-II (4%). Other devices (Waves, Airdrive HMP, CMP-X08, Kidney Assist Transport) were each used in a single study; device type was unspecified in 6% of studies.

Devices employed either roller or centrifugal pumps. Most were pressure-controlled (typically 30 mmHg; range: 15-60 mmHg), with only Gambro PF-3B and Waves using flow control. Several studies adjusted perfusion pressure based on RR or flow targets (Table S5). The most commonly used perfusion solution was University of Wisconsin Machine Perfusion Solution (UW-MPS or Belzer-MPS), reported in 29 studies (59%). Occasional use of UW-Gluconate and cryoprecipitated human plasma was reported in 2 studies each (4%). In 12 studies (24%), the perfusion solution was not specified (Table S6). Additives were reported in 13 studies, including osmotic diuretics like mannitol;^5,8,64,66,77,81^, antibiotics like penicillin,^5,49,59^ insulin,^5,49,59,83^ dexamethasone,^5,49,59,83^ and vasodilators^71,83^ (Table S6). Oxygenation was described in six studies (12%).21,28,29,31,60,63

To assess evolving practices, we performed a subanalysis based on the introduction of the LifePort device around 2000. Pre-2000 studies (n=3) used MOX 100 or Gambro PF-3b and reported longer perfusion durations (up to 20 hours).^76,81–83^ Although some of these studies included DCD donors, only one contributed extractable RR data, and this involved a DBD donor group (Fig S4). Post-2000 studies (n=45) increasingly adopted Lifeport and RM3, although substantial protocol heterogeneity persisted, particularly in pump control strategies despite more consistent use of standardized perfusion pressures (Table S5). Although RR data could not be extracted from 7 post-2000 studies, the remaining data span both DBD and DCD donors (Fig. S4). Recent studies (post-2020) reported shorter warm ischemia times and a greater focus on RR determinants outcomes.

### Renal Resistance Trajectories During Perfusion

The typical RR curve showed a rapid initial decline followed by gradual stabilization within five hours Fig. 2). Initial RR values ranged from 0.30 to 3.50 mmHg/mL/min, decreasing to 0.17-1.50 mmHg/mL/min. Due to heterogeneity in units, reporting times points, risk of bias, meta-analysis of RR evolution was not feasible.

**Fig 2.**
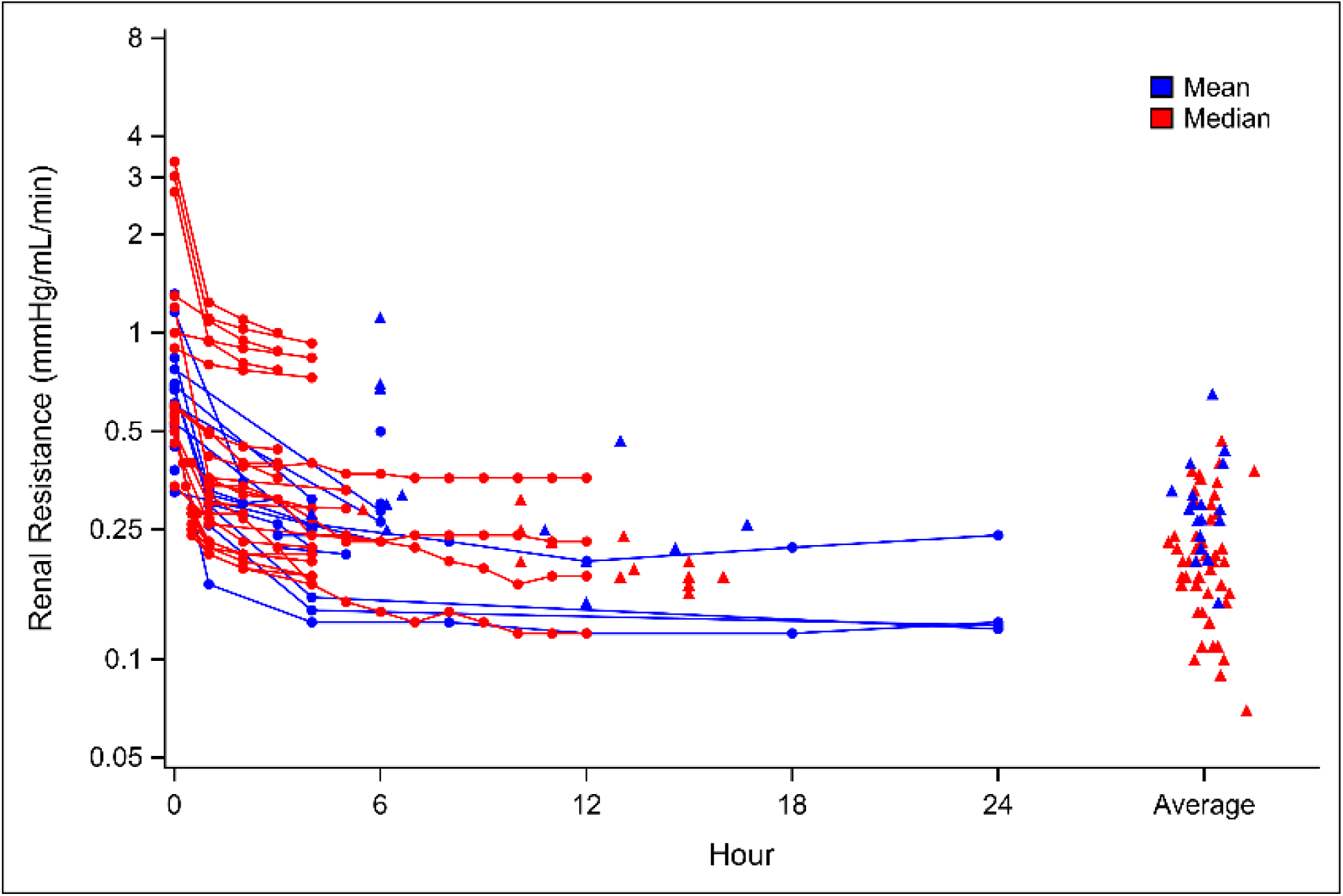
Data visualization of measured RR over the course of perfusion in the primary study-reports. Each data point represents a mean or median RR value at a specific time point or the end of perfusion (solid triangles), the latter at the mean or median time of end of perfusion. For studies with averaged RR values, a triangle is placed at the far right of the x-axis (using jitter to distinguish the studies). Solid lines represent studies contributing information on multiple time points.

As Lifeport was the most frequently used device, we compared RR-trajectories between Lifeport and other devices (Fig. S2). Lifeport initial RR values ranged from 0.30 to 1.20 mmHg/mL/min, stabilizing between 0.17 and 0.40 mmHg/mL/min within four hours. Other devices showed broader ranges: initial RR values ranged from 0.30 to 3.50 mmHg/mL/min and plateau values between 0.19 and 1.50 mmHg/mL/min. One data set involving kidneys with Karpinski scores between 5-7 showed unusually high initial RR (2.71-3.35 mmHg/ml/min).^28^ Studies using the LPS-II device reported implausibly high RR values (>100 mmHg/mL/min/g); these were excluded after failed attempts to clarify with the authors.^60,77,84,85^

### Determinants of renal resistance

Ten studies (20%) assessed RR determinants, which were grouped into histological features, donor characteristics, and perfusion additives (Table 1). Due to inconsistent reporting, diverse variables, and small sample sizes, meta-analysis was not performed.

**Table 1.**
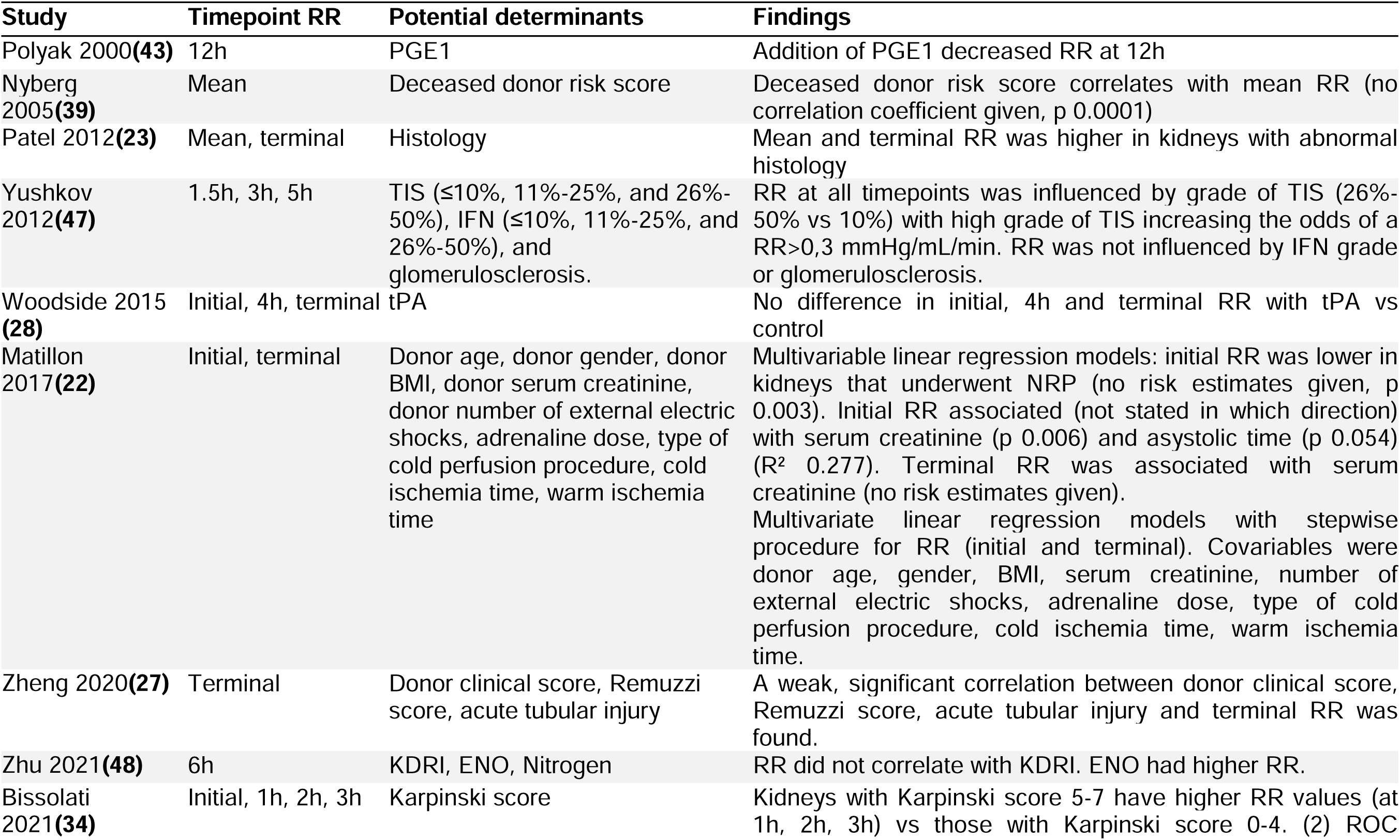
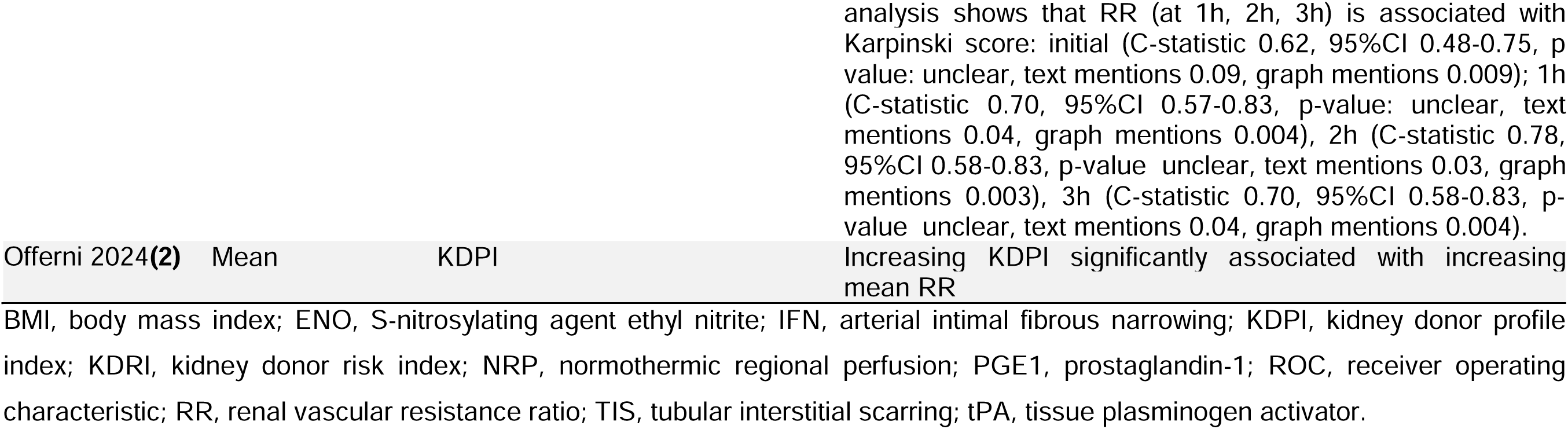
Table renal resistance determinants.

Histological findings such as Karpinski score, tubular interstitial scarring, and glomerulosclerosis were variably associated with elevated RR (Table S7).^28,30,55,56^ Bissolati et al reported that kidneys with Karpinski scores 5 to 7 had significantly higher RR early after the start of perfusion, with a cut-off RR distinguishing score groups. No specific histological components (glomerulosclerosis, fibrosis, or arterial narrowing) were identified as primary contributors.^28^ Patel et al. found a weak correlation between RR and glomerulosclerosis or hyaline arteriosclerosis.^56^ Yushkov et al. reported that tubular interstitial scarring was significantly associated with elevated RR.^55^ Zheng et al. found that the Remuzzi score correlated weakly with RR, while acute tubular injury was predictive of RR but not associated with RR.^30^

Donor characteristics and risk scores showed mixed associations: some studies reported weak correlations, while others found none. For example, Matillon et al. found that donor characteristics such as serum creatinine and asystole time were independently associated with initial RR.^45^ Only serum creatinine was independently associated with terminal RR. The findings on the association between donor risk scores and RR were also heterogeneous, with results ranging from a weak positive correlation between higher risk scores and RR,^2,30^ to no association,^26^ and an association that was not further specified.^64^ Visual inspection of RR-trajectories over time showed no consistent clustering of RR values that clearly distinguished donor types across studies or timepoints (Fig S3, S4).

Three studies assessed perfusion additives as RR determinants. Two vasodilators were studies: prostaglandin-1 (PGE1) significantly decreased RR at 12h,^71^ while adding ethyl nitrite (ENO), an S-nitrosylating agent, was associated with higher RR at 6h.^26^ Tissue plasminogen activator (tPA) had no measurable effect.^49^

### Renal Resistance and Transplant Outcomes

Twenty studies (40%) examined the association between RR and transplant outcomes, grouped as early graft function (16/49, 32%), graft survival (6/49, 12%), and kidney discard (2/49, 4%) (Table 2).

**Table 2.**
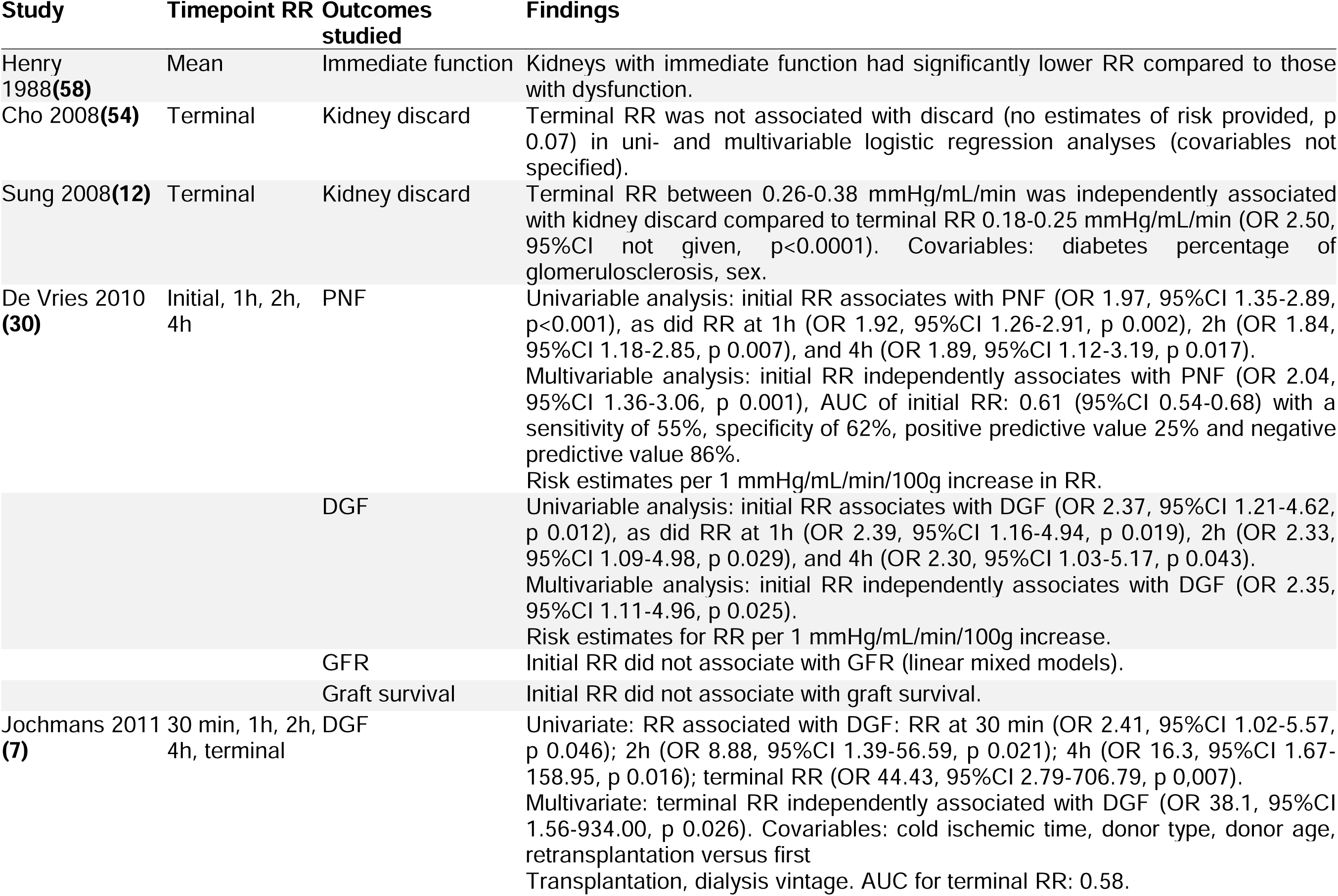
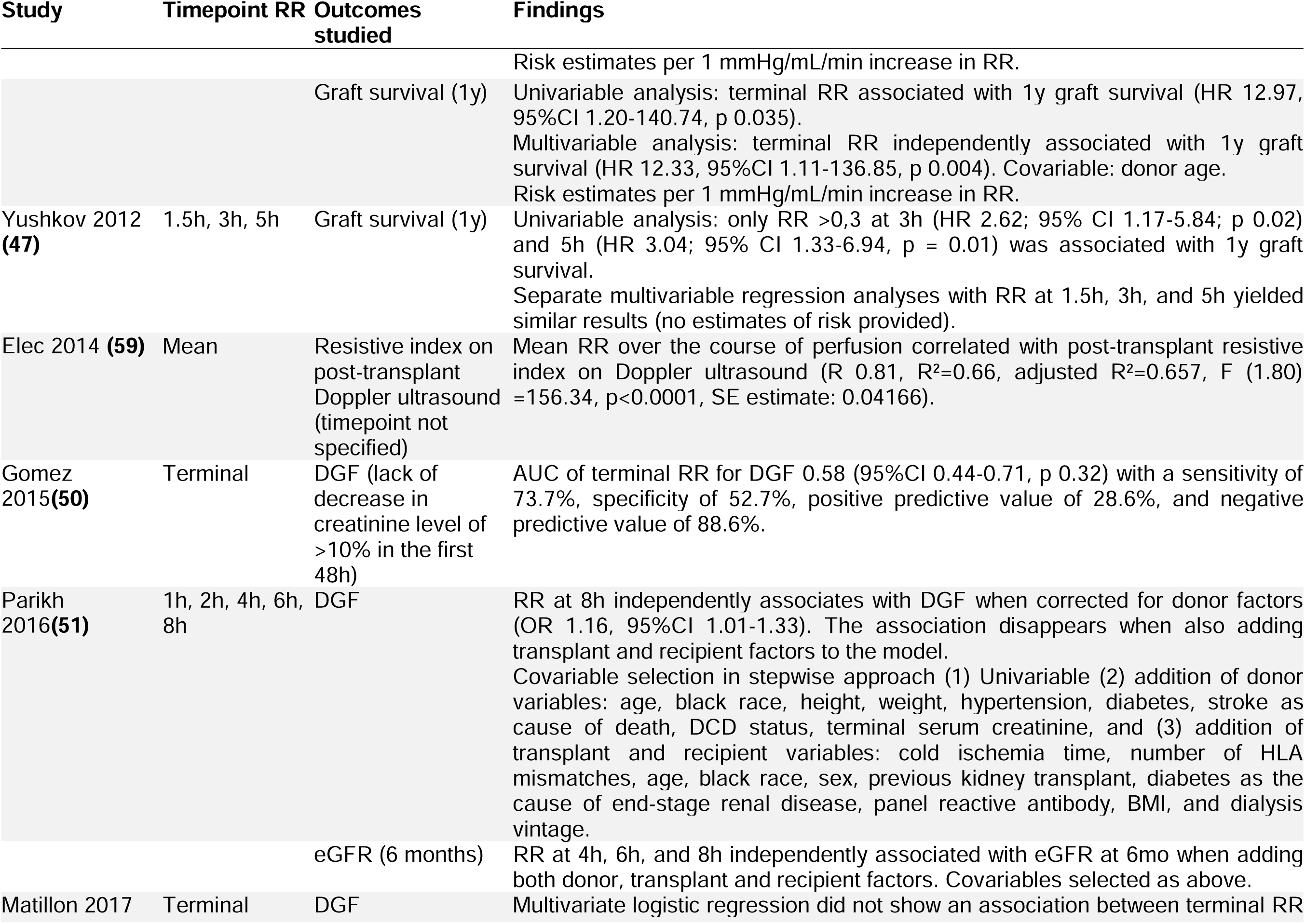
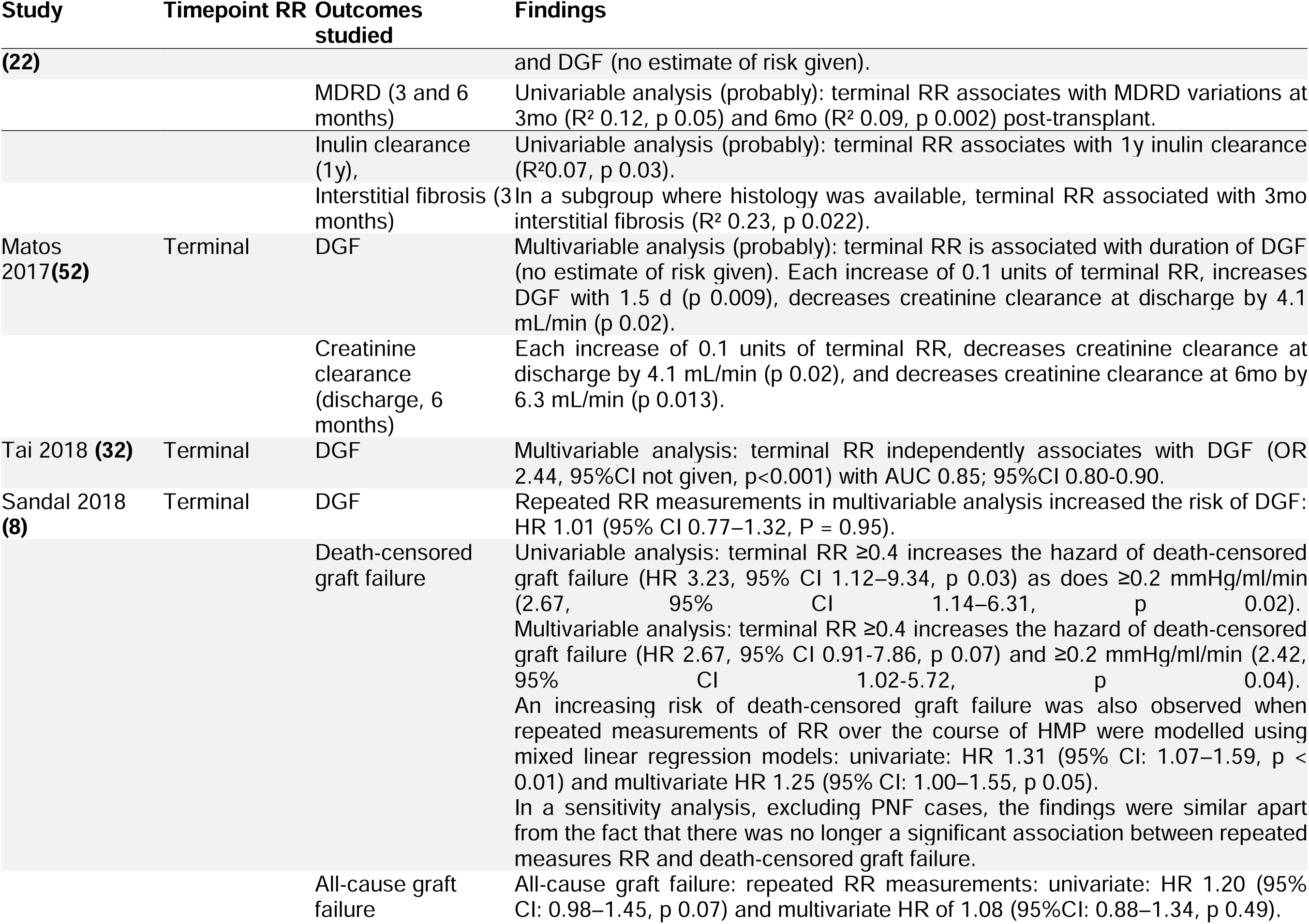
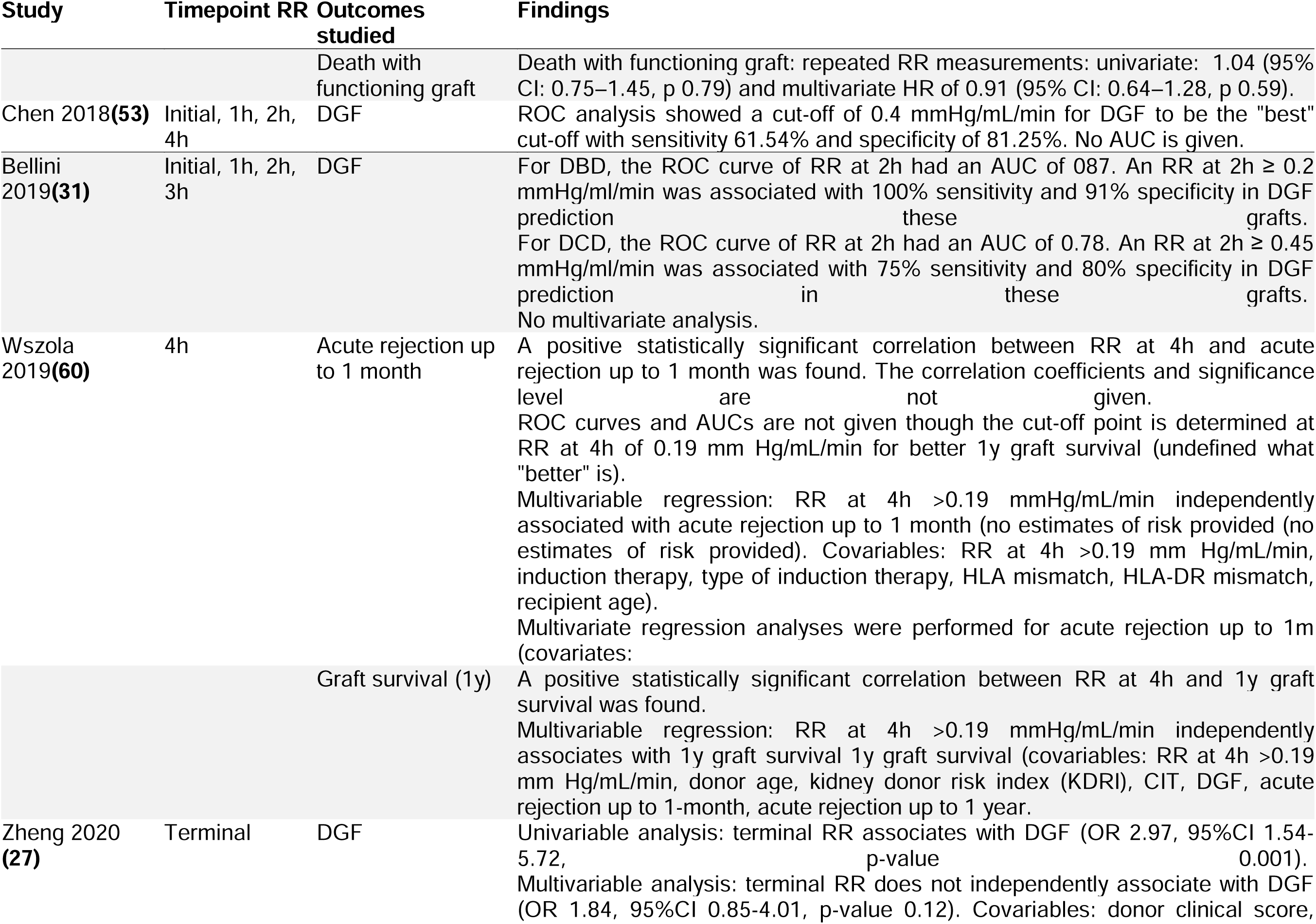
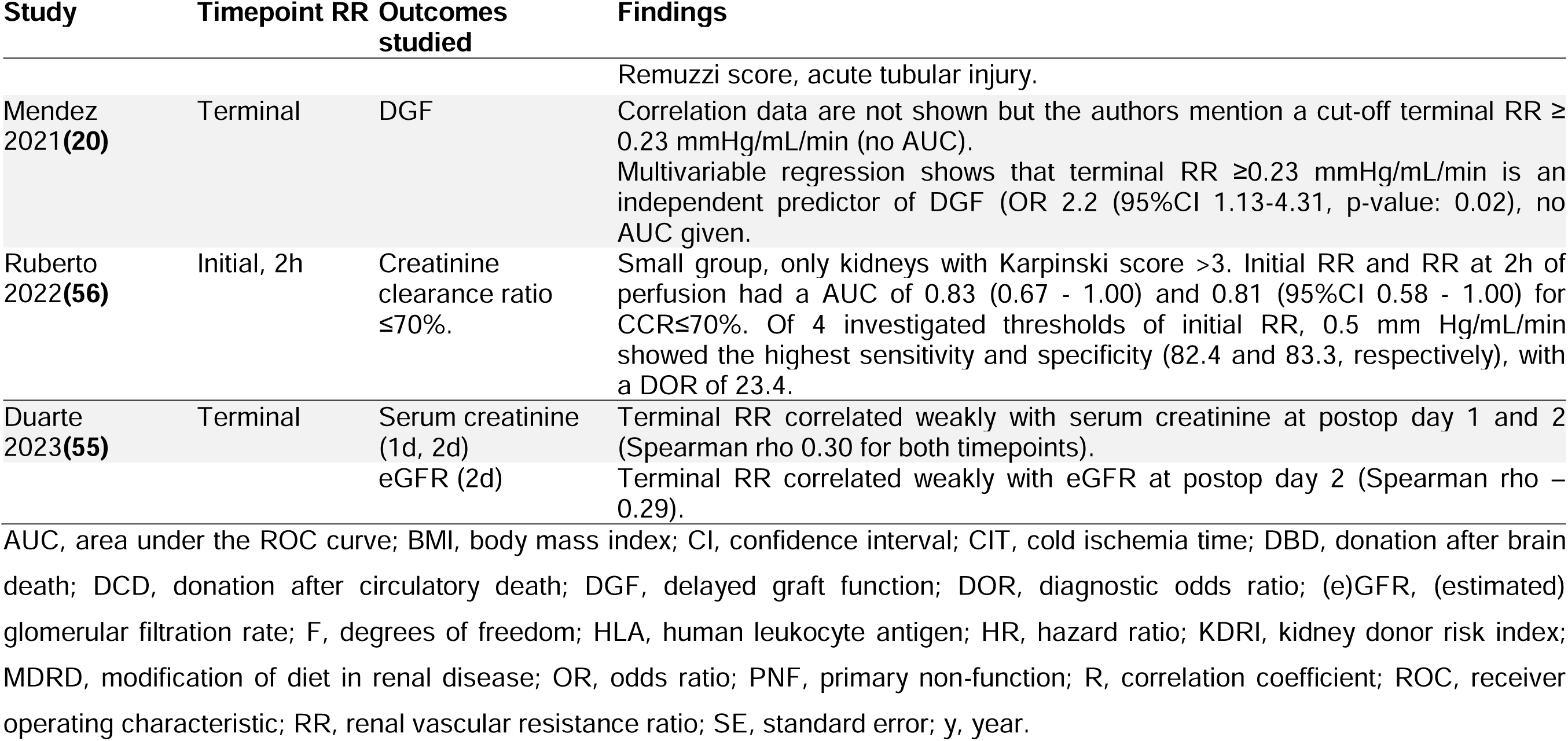
Table outcome associations.

Eleven studies (22%) linked higher RR (typically terminal RR) with DGF, while one did not find this association.^8,27,30,33,37,39,45–47,51,57,59^ The time points at which RR was measured varied widely, from terminal RR8,12,20,27,30,37,45,46,51,62,87 to multiple time points during perfusion.20,22,33,39,47,55,57 A meta-analysis of three studies confirmed a significant univariable association between terminal RR (in mmHg/ml/min) and DGF ([pooled OR 2.49 (95% CI 1.49-4.18), I^2^ 58%] (Fig 3).^23,37,57^ Sensitivity analysis confirmed the robustness of our findings (Fig. S4). Excluding Jochmans et al.^88^, which reported an extreme OR (44.43), had minimal effect on the pooled estimate (OR = 2.43, 95% CI: 0.35–137.00), indicating that it did not disproportionately influence the results.

**Fig 3.**
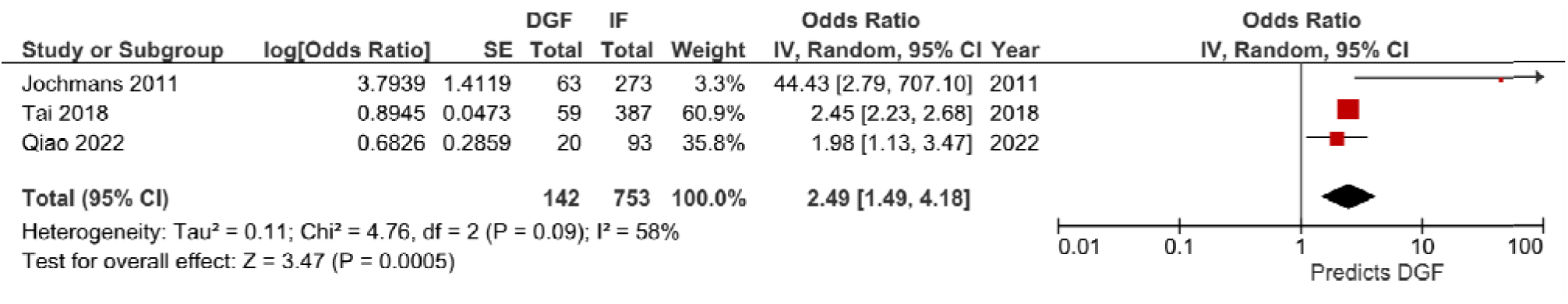
Forest plot of univariate associations between terminal renal resistance and delayed graft function. CI, confidence interval; DGF, delayed graft function; IF, immediate function; IV; inverse variance; OR, odds ratio; SE, standard error

However, excluding Tai et al.,^37^ which had the highest study weight (60.9%), resulted in a pooled OR of 6.90 (95% CI: 0.35-137), with a wider confidence interval, highlighting its strong influence on the precision of the pooled estimate. The pooled univariable AUC for terminal RR predicting DGF was 0.77 (95% CI 0.66-0.88). However, in reported multivariable models, a lower predictive accuracy was described (AUC 0.58-0.74).^33,57,59^

Several studies proposed RR thresholds for predicting outcomes, though only one was robust (Tables S8, S9). Most thresholds lacked validation and were based on limited statistical rigor.

Four studies assessed immediate function, with lower RR values generally associated with better outcomes.^39^ Five studies linked RR with graft survival.^27^

Two studies assessed kidney discard. One found no association, another showed higher terminal RR (0.26-0.38 mmHg/mL/min) increased discard likelihood.^12,62^

## Discussion

This study systematically evaluated the role of RR during HMP, identifying key limitations in how it is measured, interpreted, and used in clinical practice. Although RR is commonly used to guide kidney acceptance, its predictive reliability remains uncertain due to inconsistent measurement practices, variability in timing, and lack of adjustment for confounding factors. The nearly four-decade span of included studies (1988–2024) reflects the evolution of HMP technology but introduces substantial challenges due to variability in devices, protocols, donor standards, and transplantation practices. Most studies were retrospective and at moderate to serious risk of bias, and their uneven geographical distribution further complicates interpretation. While recent studies have trended towards standardization, heterogeneity remains a central limitation. These findings highlight the need for contemporary, standardized research, as earlier data may not reflect current clinical practice.

By aggregating all available RR data, we identified a distinct trajectory characterized by a steep initial decline followed by a gradual levelling off to approach a plateau within the first 4-5 hours of perfusion. However, most studies assess RR values at a single time point – typically terminal – neglecting the valuable information embedded in the full trajectory. Intermediate RR measurements have received limited attention, yet may hold key predictive value. The full trajectory of RR has rarely been systematically investigated, underscoring the need for dedicated studies to explore dynamic RR behaviour throughout perfusion.

While RR is frequently used in clinical practice, our findings suggest its standalone predictive power is modest. Our meta-analysis confirmed that higher RR values are associated with an increased risk of DGF, with a pooled OR of 2.49 and an AUC of 0.77. Sensitivity analyses showed that this association remained robust despite the exclusion of individual influential studies. However, predictive accuracy declined in reported multivariable models, with AUCs ranging from 0.58 to 0.74, emphasizing the limitations of RR as a standalone parameter to discard kidneys.

Numerous studies proposed RR thresholds for predicting transplant outcomes, yet these varied widely and were rarely validated. Only two studies met the criteria for high methodological robustness. The absence of a clear inflection point suggests an incremental, rather than binary, relationship between RR and post-transplant outcomes. While thresholds were derived using ROC analysis, variability in device settings, perfusion protocols, and measurement time points preclude the definition of a universal cut-off. Moreover, existing studies have not systematically applied likelihood ratios, dose-response modeling, or similar approaches to explore threshold behavior. Given these limitations, we refrained from conducting a poole threshold analysis. Future research should move beyond simplistic thresholds toward risk stratification models that incorporate RR alongside other clinical parameters.

Identifying factors that influence RR is essential to interpreting its clinical utility. RR is directly dependent on perfusion pressure and flow, which are influenced by device setting, pump type, perfusate viscosity, temperature, and additives affecting the kidney’s vascular bed. Our findings suggest that LifePort devices tend to produce lower and more consistent RR stabilization compared to other devices. However, as LifePort was the most commonly used device in the included studies, this observed consistency may partly reflect greater data availability rather than intrinsic device characteristics. Additionally, substantial overlap in RR values across devices complicates direct comparisons and limits the establishment of device-independent thresholds. Surprisingly, renal weight—a biologically plausible determinant of RR—has not been explicitly examined, representing a significant gap. Perfusate additives such as vasodilators may influence RR, but research is limited and protocols vary widely. Donor characteristics such as donor type may also affect RR, though variation within donor types and substantial overlap limit their interpretive value. Similarly, correlations between histological features and RR were inconsistent across studies. While some suggested associations with tubular scarring or fibrosis, findings were neither uniform nor clearly linked to transplant outcomes. A lack of standardized histological assessment and absence of data on discarded kidneys further limit these insights.

Given the substantial variability in RR measurement timing, device settings, thresholds, and reporting practices, there is an urgent need for standardization. We propose a core dataset (Table S10) to support harmonized data collection across transplant centers, incorporating essential perfusion parameters as well as donor, recipient, and outcome variables. To address these inconsistencies and improve comparability, we propose a core dataset designed to harmonize RR-related data collection across transplant centers. International initiatives, such as the Registries Platform of the European Society for Organ Transplantation (ESOT), are aligned with these goals and may facilitate broader adoption.^89^ Harmonizing measurement time points, incorporating device-specific RR adjustments, and standardized variables such as perfusate composition, temperature, and flow would enhance the reproducibility – and possibly the clinical utility – of RR.

The extended time span covered in this review introduces considerable variability, emphasizing the need for era-specific and device-specific subgroup analyses that were unable to be conducted with the current data. Future studies should explore how evolving technologies, clinical protocols, and donor characteristics have influenced RR reporting and interpretation. A significant limitation identified in our review is the lack of data on discarded kidneys. Because these grafts are often excluded from analysis, we lack insight into how RR thresholds may contribute to inappropriate discards. This selection bias is a critical blind spot in the current literature.

In conclusion, RR remains a widely used but inconsistently applied marker in kidney transplantation. While it shows modest predictive potential for outcomes like DGF, its clinical application is limited by heterogeneity in measurement and interpretation. Future research should prioritize large-scale registry data and advanced analytics – such as machine learning – to build robust, multivariable risk models that integrate RR with other key indications. Until such tools are validated, we strongly advise against using RR thresholds alone to guide kidney acceptance decisions. Instead, RR should be incorporated into comprehensive, evidence-based frameworks aimed at optimizing transplant outcomes and minimizing unnecessary organ discard.

## Data Availability

All data that support the findings of this study are openly available in RDR, the KU Leuvens repository at https://doi.org/10.48804/YG4LXD

## Acknowledgments

We thank Veerle Heedfeld and Tine Wylin for their help with data extraction, quality assessment, and table preparation. We also thank Thomas Vandendriessche, Chayenne Van Meel, and Krizia Tuand, biomedical reference librarians of the KU Leuven Libraries—2Bergen—Learning Centre Désiré Collen (Leuven, Belgium) for their help in conducting the systematic literature search.

## Supporting information

Additional supporting information may be found online in the Supporting Information section.

### Supplemental Digital Content

#### Supplementary Figures

Fig S1 Study flow diagram.

Fig S2 Data visualization of measured RR over the course of perfusion in the primary study-reports split by perfusion device.

Fig S3 Data visualization of measured RR over the course of perfusion in the primary study-reports split by era.

Fig S4 Data visualization of measured RR over the course of perfusion in the primary study-reports split by donor type.

Fig S5 Quality assessment ROBINS I tool.

Fig S6 Leave-One-Out sensitivity analysis of terminal renal resistance and delayed graft function (DGF) using a random-effects model.

#### Supplementary Tables

Table S1: Primary study reports organized by allocation region.

Table S2: Overview of articles by allocation region, subgroups, donor type, donors, donors’ age, warm ischemia time, and perfusion time.

Table S3: Overview of primary study reports that selected kidneys based on perfusion parameters introducing a selection bias.

Table S4: Perfusion devices.

Table S5: Pump control adaptation. Table S6: Perfusion characteristics.

Table S7: Histological findings and their association with renal resistance (RR) during hypothermic machine perfusion.

Table S8: Renal Resistance threshold studies: summary of findings and robustness grading.

Table S9: Criteria-based grading of study robustness in RR threshold research.

Table S10: Essential data elements for transplant registries integrating machine perfusion.

## Abbreviations

AH: abnormal histology
AP: anterograde perfusion
AUC: area under the curve
BMI: body mass index
CI: confidence interval
CIT: cold ischemia time
DBD: donation after brain death
DCD: donation after circulatory death
DGF: delayed graft function
DKT: double kidney transplantation
DOR: diagnostic odds ratio
ECD: expanded criteria donors
eGFR: estimated glomerular filtration rate
ENO: S-nitrosylating agent ethyl nitrite
F: degrees of freedom
GFR: glomerular filtration rate
GP: good perfusion
HLA: human leukocyte antigen
HR: hazard ratio
HTK: histidine-tryptophane-ketoglutarate
IF: immediate function
IFN: arterial intimal fibrous narrowing
IGL: Institut Georges Lopez
IV: inverse variance
KDPI: kidney donor profile index
KDRI: kidney donor risk index
MDRD: modificaton of diet in renal disease
MP: marginal perfusion
NG: not given
NH: normal histology
NRP: normothermic regional perfusion
OR: odds ratio
PGE1: prostaglandin-1
PNF: primary non function
R: correlation coefficient
ROC: receiver operating characteristic
RP: retrograde perfusion
RR: renal resistance
SE: standard error
SKT: single kidney transplantation
TIS: tubular interstitial scarring
tPA: tissue plasminogen activator
UW: University of Wisconsin
y: year

