## Supplementary Appendix for "Renal Resistance During Hypothermic Machine Perfusion: A Scoping Review of Variability and Determinants, with a Meta-Analysis of Predictive Value for Transplant Outcomes"

**Author affiliations**

Laurence Verstraeten: 0000-0002-1323-2105

Steffen Fieuws: 0000-0002-6875-8366

Ina Jochmans: 0000-0003-4592-2810

### Appendices

##### Appendix S1 Search strategies

***PUBMED***

"Kidney"[mesh] OR "kidney*"[tiab] OR “renal*”[tiab] OR ”kidney transplant*”[tiab] OR ”renal transplant*”[tiab] OR ”kidney graft*”[tiab] OR ”renal graft*”[tiab]

AND

"Vascular resistance"[Mesh] OR "resist*"[tiab]

AND

“Perfusion"[mesh] OR “perfus*”[tiab] OR “Organ Preservation"[Mesh]

***EMBASE***

"Kidney"/exp OR "kidney*":ti,ab,kw OR “renal*”:ti,ab,kw OR ”kidney transplant*”:ti,ab,kw OR ”renal transplant*”:ti,ab,kw OR” kidney graft*”:ti,ab,kw OR” renal graft*”:ti,ab,kw

AND

"Vascular resistance"/exp OR "resist*":ti,ab,kw

AND

'perfusion'/exp OR 'perfus*':ti,ab,kw OR 'organ preservation'/exp

***WEB OF SCIENCE***

TS=("Kidney" OR "kidneys" OR "renal" OR "renal*" OR "Kidney transplantation" OR "kidney

transplant*" OR "renal transplantation" OR "renal transplant*" OR "kidney graft*" OR "renal graft*")

AND

TS= ("resistance" OR "resistance*" OR " resist*")

AND

TS= ("Perfusion" OR "Organ preservation" OR “perfus*”)

***COCHRANE LIBRARY***

"kidney" OR "kidney transplantation"OR ("kidney" OR "kidneys" OR "renal" OR "kidney NEXT transplant" OR "renal NEXT transplant" OR "kidney NEXT graft" OR "renal NEXT graft")

AND

"perfusion"	OR	"organ preservation"	OR	("perfusion" OR "organ preservation" OR "perfus")

AND

"Vascular resistance" OR ("vascular NEXT resistance" OR "resistance" OR "resist" OR "resistances" OR "resistant" OR "resistive" OR "resist")

The results of all database searches were uploaded in Rayyan (insert citation). The resulting articles were screened for duplicates using the “Find Duplicates” tool in Rayyan and for language using Rayyan “Language” filters.

##### Appendix S2 Inclusion and exclusion criteria for studies

**INCLUSION CRITERIA**

- Language: English, Dutch, French or Spanish.
- Research articles: original research articles, systematic or scoping reviews.
- Years considered: no limits
- Content:
  - Study must be performed with human kidneys
  - Kidneys must be isolated from the body by hypothermic (< 12°C) ex-situ machine
  - perfusion techniques
  - Studies should report on perfusion parameters during hypothermic kidney
  - perfusion
- Full text available (freely online or via KU Leuven Association)

**EXCLUSION CRITERIA**

- Language: other than English, Dutch, French or Spanish.
- Study type: non-original research articles (review articles, letter to editor, conference abstracts,
- editorials etc.), qualitative research.
- Content:
  - Studies not using human kidneys
  - Only organs other than kidneys were examined
  - Studies where perfusion was not hypothermic perfusion (≥ 12°C)
  - Studies not reporting on perfusion parameters during hypothermic kidney perfusion
- No full text available

##### Appendix S3 Selection of primary study report

Several articles might report on the same study or on repeated data sets. Such duplicate publication can take various forms, ranging from identical manuscripts to reports describing different outcomes of the study or results at different time points ^1^. As duplicate publication and repeated data sets can introduce substantial biases if studies are inadvertently included more than once in a meta-analysis ^2^, multiple reports of the same study were linked and not considered as multiple studies. The following methodology was used to identify duplicate publication or repeated data sets: (partial) repetition of data sets between articles was assessed based on country and author affiliations, intervention groups and specifics of the intervention, the recruiting period, date and duration of the study. When overlap between reported data was suspected, the authors were asked whether they could confirm the suspected overlap. When no reply was received, the suspected overlap was assumed to be present. When (partial) repeated data sets reported in different articles was identified, multiple reports of the same study were collated, so that each study, rather than each report, is the unit of interest in this review. A primary study report was chosen for each study based on the number of participants in the intervention group of interest and the date of publication. Since other reports may contain additional outcome measures of importance, other comparator groups or other valuable information, these reports were not discarded but relevant information was extracted from these papers and these data are mentioned or used in quantitative analyses when relevant.

##### Appendix S4 Data visualization

Renal vascular resistance data were extracted from primary study reports. Point estimates were determined from available graphs, using WebPlotDigitizer v.4.3 (Ankit Rohatgi, California, USA), unless the necessary data points were reported ^3,4^. Each data point represents mean or median RR values at specific time points or, in cases where RR was measured as terminal RR, the length of the perfusion was used. When no precise length of perfusion was reported and if the perfusion was continuous and the cold ischemia time was provided, the cold ischemia time was assumed to represent the duration of perfusion. Some papers only presented RR values averaged over the entire perfusion. All “average” values are presented on the far right of the x-axis, irrespective if the perfusion time was mentioned in the paper. To avoid confusion with the “terminal RR” values (i.e. those at the end of perfusion), the RR values averaged over the length of perfusion with a known mean/median time were not plotted at their mean/median time, they are only plotted once (i.e. at the far right of the X-axis).

For studies reporting RR in units of mmHg/mL/min/100 g, where kidney weight was provided, the values were converted to mmHg/mL/min by using the mean kidney weight reported in the paper. In cases where the mean or median kidney weight was not provided, an estimated average kidney weight of 141 g was used, based on a population-wide average derived from typical anatomical references ^5^. This estimation was calculated from the following: Male kidney: 125–170 g; Female kidney: 115–155 g. The population-wide average kidney weight, assuming equal male and female distribution, was estimated to be 141.25 g. The standard deviation for this population-wide kidney weight was assumed to be approximately 10.63 g.

Data variability is not represented in this figure, as the data points are based on the mean or median values extracted from the literature. For visualization, jitter has been applied to the timing of the values, which represent either the full perfusion average or measurements at key time points.

##### Appendix S5 Clinical heterogeneity and grouping of studies for meta-analyses

Clinical heterogeneity was assessed by extracting baseline donor and recipient demographics. Data pooling was considered although clinical heterogeneity was considered likely as the target patient population for hypothermic kidney perfusion is heterogeneous. Studies were grouped regardless of the donor type when at least 3 studies reported on the outcome of interest on a univariable level. Grouping of studies that reported on the outcome of interest at multivariable level was considered though abandoned as the few studies that did report multivariable results corrected for different covariables in their analyses.

##### Appendix S6 Robustness Grading Criteria

The methodological quality of studies proposing renal resistance (RR) thresholds in relation to post-transplant outcomes (e.g., DGF, PNF, graft function, survival) was evaluated according to the criteria below. The robustness level reflects how appropriate a study is as evidence to guide decisions—especially when thresholds may be used to accept or discard donor kidneys.

**High Robustness**

A study is graded as *High Robustness* only if all of the following criteria are met:

1. Threshold derivation: The RR threshold is derived using appropriate statistical methods (e.g., ROC curve, Youden Index).
2. Multivariable adjustment: Analysis includes adjustment for relevant confounders such as donor age, cold ischemia time, and KDPI.
3. Predictive performance: The study reports predictive metrics such as AUC, sensitivity/specificity, PPV/NPV, or odds/hazard ratios with confidence intervals.
4. No selection bias: The study includes all relevant grafts without discarding organs based on RR/RI prior to analysis.
5. Adequate sample size: The number of transplanted cases is at least 150.
6. Validation: The threshold is validated internally (e.g., cross-validation, bootstrapping) or externally in an independent cohort.

**Moderate Robustness**

A study is graded as *Moderate Robustness* when all of the following are true:

1. Statistical association tested: RR/RI is statistically associated with outcomes using univariate or basic statistical tests (e.g., t-test, ANOVA, log-rank).
2. Threshold plausible but not derived: A cutoff is used that is plausible or cited from prior literature but not derived from the study’s own data.
3. No or limited adjustment: There is no multivariable analysis, or it is incomplete.
4. Limited predictive performance: Predictive performance is either not reported, or the AUC is < 0.70 or lacks interpretation.
5. Sample size between 50 and 149 transplanted cases, or sample size is unclear.

**Low Robustness**

A study is graded as *Low Robustness* if two or more of the following apply:

- The RR threshold is arbitrary, borrowed without justification, or based solely on anecdotal practice.
- No statistical association is tested between RR and clinical outcomes.
- No multivariable adjustment is performed.
- No predictive metrics (e.g., AUC, OR/HR) are reported.
- The sample size is fewer than 50 transplant cases.
- Selection bias is likely or explicitly present (e.g., grafts excluded from analysis due to high RR).

### Supplementary Figures

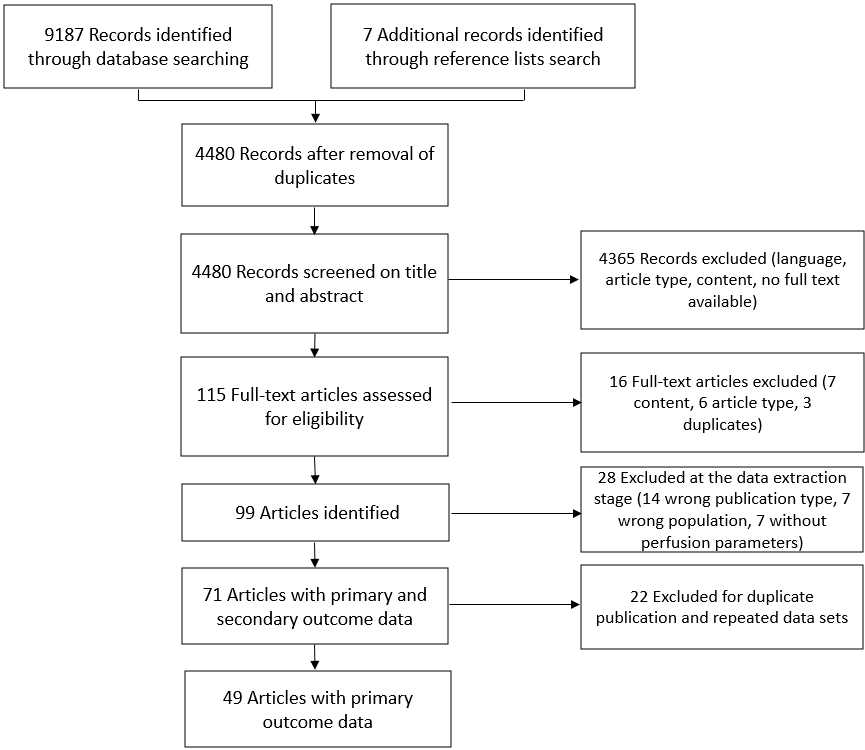

*Fig S1 Study flow diagram*

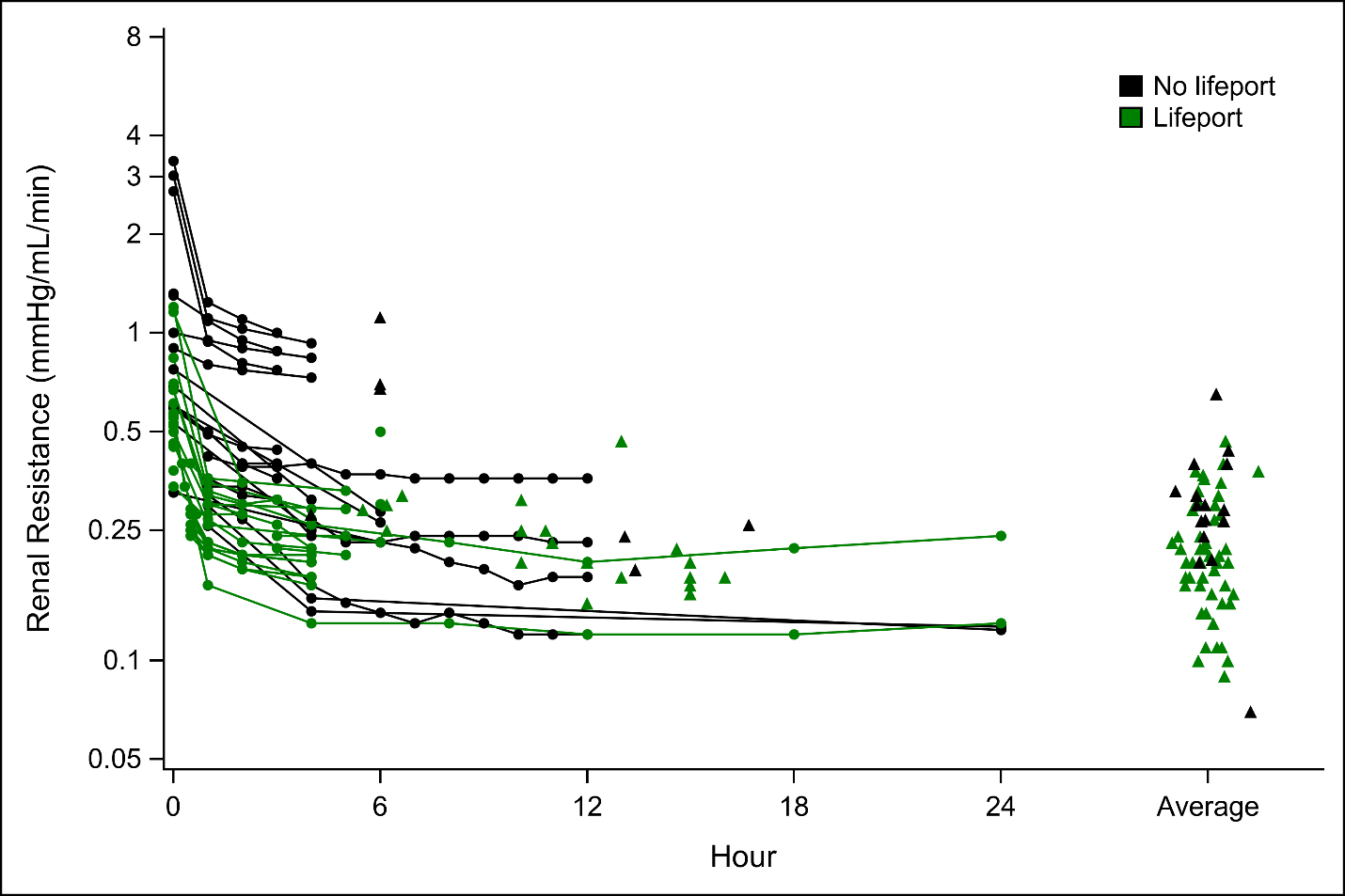

*Fig S2 Data visualization of measured RR over the course of perfusion in the primary study-reports split by perfusion device.*

*Each data point represents a mean or median RR value at a specific time point or the end of perfusion (solid triangles), the latter at the mean or median time of end of perfusion. For studies with averaged RR values, a triangle is placed at the far right of the x-axis (using jitter to distinguish the studies). Solid lines represent studies contributing information on multiple time points.*

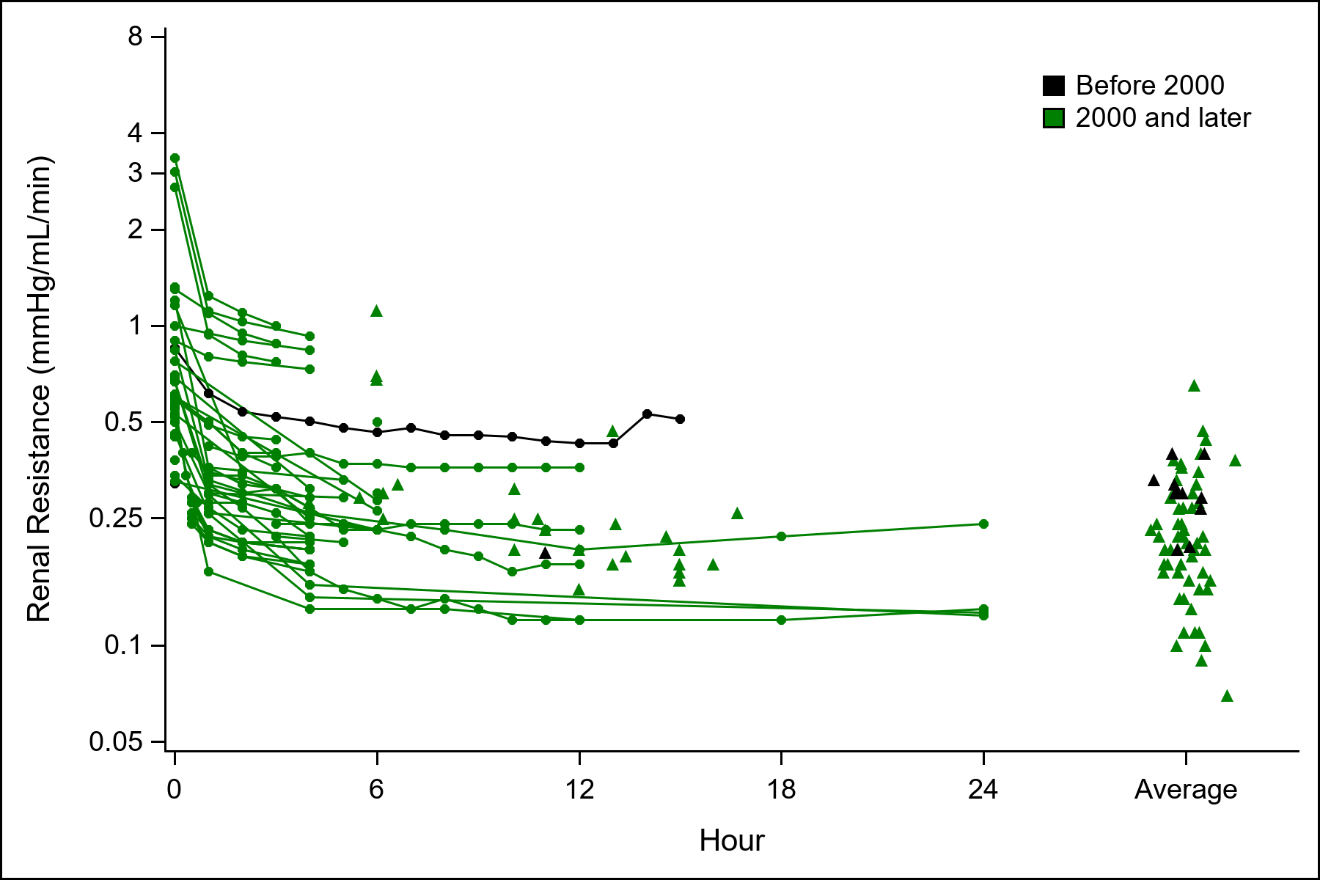

*Fig S3 Data visualization of measured RR over the course of perfusion in the primary study-reports split by era.*

*Each data point represents a mean or median RR value at a specific time point or the end of perfusion (solid triangles), the latter at the mean or median time of end of perfusion. For studies with averaged RR values, a triangle is placed at the far right of the x-axis (using jitter to distinguish the studies). Solid lines represent studies contributing information on multiple time points.*

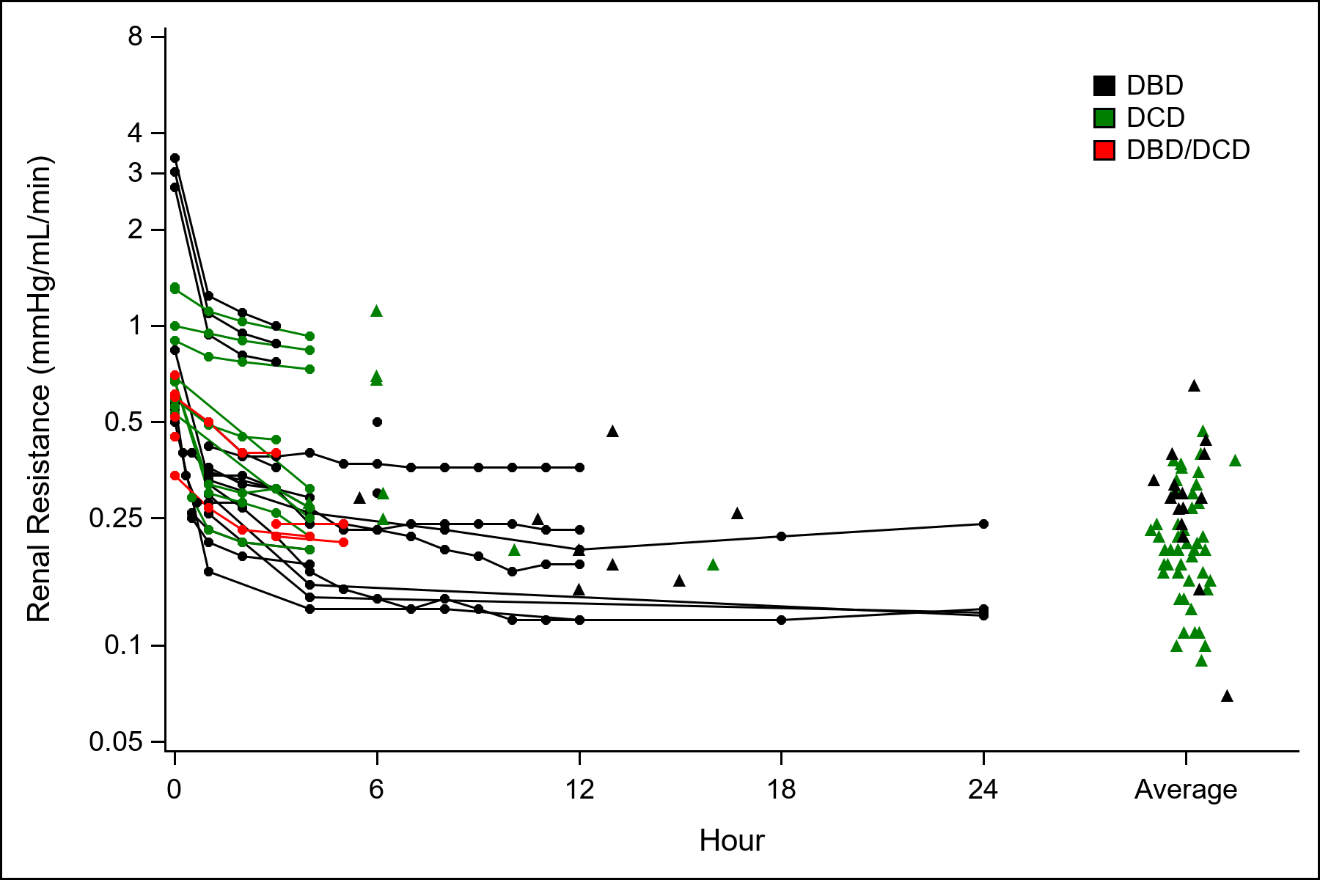

*Fig S4 Data visualization of measured RR over the course of perfusion in the primary study-reports split by donor type.*

*Each data point represents a mean or median RR value at a specific time point or the end of perfusion (solid triangles), the latter at the mean or median time of end of perfusion. For studies with averaged RR values, a triangle is placed at the far right of the x-axis (using jitter to distinguish the studies). Solid lines represent studies contributing information on multiple time points.*

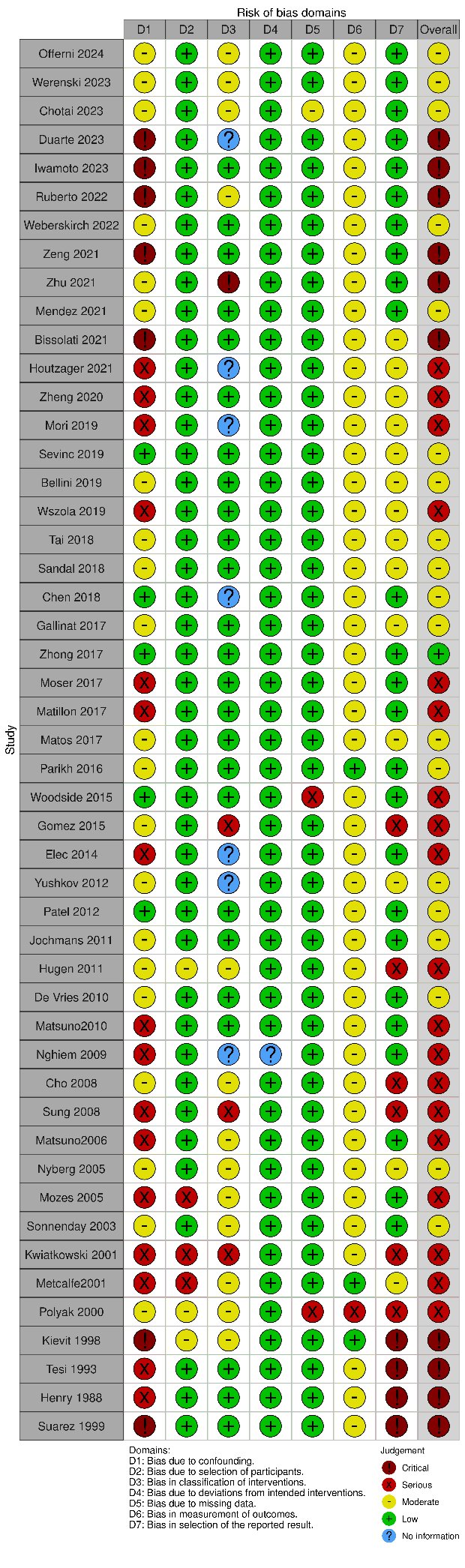
*Fig S5 Quality assessment ROBINS I tool*

**A**
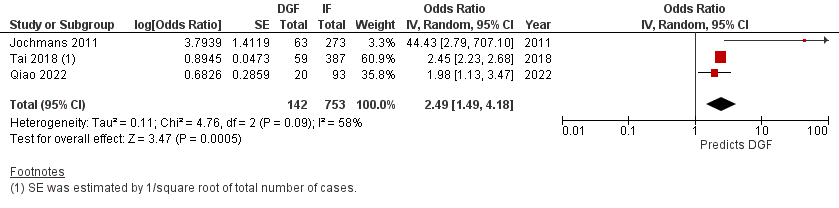

**B**
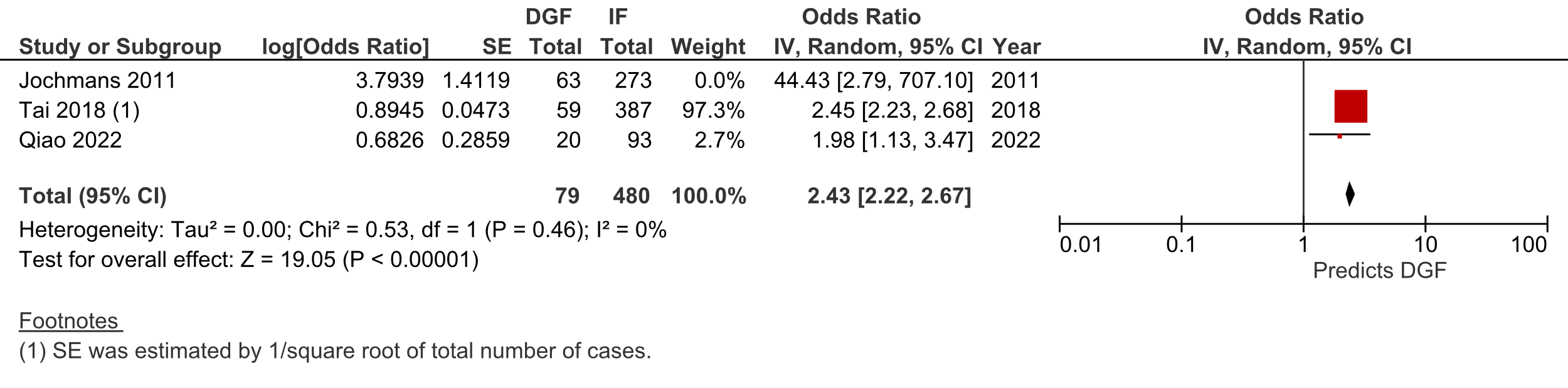

**C**
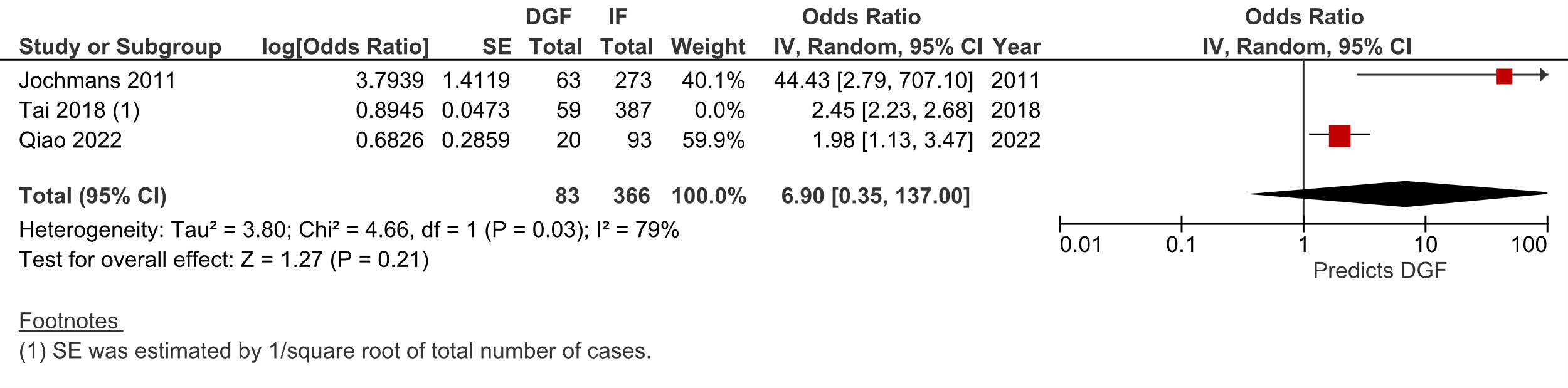

*Fig S6 Leave-One-Out sensitivity analysis of terminal renal resistance and delayed graft function (DGF) using a random-effects model*

*(A) Forest plot of the full meta-analysis, including all three studies (Jochmans 2011, Tai 2018, and Qiao 2022) assessing the association between terminal renal resistance (RR) and DGF. The pooled odds ratio (OR) is 2.49 (95% CI: 1.49–4.18), with moderate heterogeneity (I² = 58%).*

*(B) Forest plot of the leave-one-out analysis excluding Jochmans 2011, which reported an extreme OR (44.43) with a wide confidence interval. The pooled estimate remains 2.43 (95% CI: 2.22–2.67), confirming that Jochmans 2011 does not substantially influence the meta-analysis results.*

*(C) Forest plot of the leave-one-out analysis excluding Tai 2018, the study with the largest weight (60.9%). The pooled OR shifts to 6.90 (95% CI: 0.35–137.00), with a wider confidence interval, highlighting that Tai 2018 plays a key role in determining the precision of the pooled estimate.*

*These sensitivity analyses confirm that the overall association between terminal RR and DGF is robust and not driven by a single study. However, the strong influence of Tai 2018 suggest that additional well-powered studies are needed to further validate this relationship and improve the generalizability of findings.*

### Supplementary Tables

*Table S1 Primary study reports organized by allocation region*

| **Primary study report** | **Secondary study reports** | **Repeated data sets assessment** |
| --- | --- | --- |
| ***Brazil*** |  |  |
| Matos 2017^6^ | - | - |
| ***Canada*** | | |
| Moser 2017^7^ | - | - |
| Sandal 2018^8^ | - | - |
| ***China*** |  |  |
| Zhong 2017^9^ | - | - |
| Chen 2018^10^ | - | - |
| Tai 2018^11^ | Ding 2018^12^ | Data from Ding 2018 (09-2012 to 08-2016) are duplicated in Tai 2018 |
| Zheng 2020^13^ | Qiao 2022^14^ | Data from Qiao 2022 (01-2019 to 08-2019) are duplicated in Zheng 2020 |
| Zeng 2021^15^ | - | - |
| ***Eurotransplant*** |  |  |
| Kievit 1998^16^ | - | - |
| De Vries 2010^17^ | Daemen 1997^18^ | Data from Daemen 1997 (07-1993 to 11-1995) are duplicated in De Vries 2010 |
| Jochmans 2011^19^ | - | - |
| Gallinat 2017^20^ | - | - |
| Houtzager 2021^21^ | - | - |
| Weberskirch 2022^22^ | - | - |
| ***France*** |  |  |
| Matillon 2017^23^ | - | - |
| ***Italy*** |  |  |
| Mori 2019^24^ | - | - |
| Bissolati 2021^25^ | Bissolati 2018^26^ | Data from Bissolati 2018 (12-2015 to 10-2017) are duplicated in Bissolati 2021 |
|  | Bissolati 2019^27^ | Data from Bissolati 2019 (01-2015 to 09-2018) are duplicated in Bissolati 2021 |
| Ruberto 2022^28^ | - | - |
| Offerni 2024^29^ | - | - |
| ***Japan*** |  |  |
| Matsuno 2006^30^ | Matsuno 1994^31^ | Data from Matsuno 1994 are strongly suspected to be duplicated in Matsuno 1996, Matsuno 1998, and Matsuno 2006 |
|  | Matsuno 1996^32^ | Data from Matsuno 1996 are strongly suspected to be duplicated in Matsuno 1998 and Matsuno 2006 |
|  | Matsuno 1998^33^ | Data from Matsuno 1998 are strongly suspected to be duplicated in Matsuno 2006 |
|  | Kozaki 2000^34^ | Data from Kozaki 2000 are strongly suspected to be duplicated in Matsuno 2006 |
| Matsuno 2010^35^ | - | - |
| Iwamoto 2023^36^ | - | - |
| ***Poland*** |  |  |
| Kwiatkowski 2001^37^ | Danielewickz 1997^38^ | Data from Danielewickz 1997 (06-1994 to 04-1997) are duplicated in Kwiatkowski 2001 |
|  | Kosieradzki 2002^39^ | Data from Kosieradzki 2002 (1994 to 1999) are duplicated in Kwiatkowski 2001 |
| Wszola 2019^40^ | Diuwe 2017^41^ | Data from Diuwe 2017 (04-2001 to 02-2014) are duplicated in Wszola 2019 |
| ***Romania*** |  |  |
| Elec 2014^42^ | - | - |
| ***Spain*** |  |  |
| Suarez 1999^43^ | - | - |
| Gomez 2015^44^ | Burgos Revilla 2015^45^ | Data from Burgos Revilla (02-2012 to 03-2014) is duplicated in Gomez 2015 |
| ***United Kingdom*** |  |  |
| Metcalfe 2001^46^ | - | - |
| Bellini 2019^47^ | - | - |
| Sevinc 2019^48^ | Sevinc 2016^49^ | Data from Sevinc 2016 (2002 to 2014) is duplicated in Sevinc 2019 |
| ***United States of America*** | |  |
| Henry 1988^50^ | - | - |
| Tesi 1993^51^ | - | - |
| Polyak 2000b^52^ | - | - |
| Polyak 2000a^53^ | Polyak 1997^54^ | Data from Polyak 1997 are duplicated in Polyak 2000a |
|  | Polyak 1999^55^  Polyak 2000b^52^ | Data from Polyak 1999 are duplicated in Polyak 2000a  Data from Polyak 2000b are duplicated in Polyak 2000a |
|  | Polyak 2000c^56^ | Data from Polyak 2000c are duplicated in Polyak 2000a |
| Sonnenday 2003^57^ | - | - |
| Mozes 2005^58^ | - | - |
| Nyberg 2005^59^ | - | - |
| Cho 2008^60^ | - | - |
| Sung 2008^61^ | - | - |
| Nghiem 2009^62^ | - | - |
| Hugen 2011^63^ | - | - |
| Patel 2012a^64^ | Patel 2012b^65^ | Data from Patel 2012b are duplicated in Patel 2012a |
| Yushkov 2012^66^ | - | - |
| Woodside 2015^67^ | - | - |
| Parikh 2016^68^ | Hall 2014^69^ | Data from Hall 2014 (05-2010 to 04-2013) are duplicated in Parikh 2016 |
|  | Doshi 2017^70^ | Data from Doshi 2017 (05-2010 to 04-2013) are duplicated in Parikh 2016 |
| Mendez 2021^71^ | - | - |
| Zhu 2021^72^ | - | - |
| Chotai 2023^73^ | - | - |
| Duarte 2023^74^ | - | - |
| Werenski 2023^75^ | - | - |

*Table S2 Overview of articles by allocation region, subgroups, donor type, donors, donors age, warm ischemia time and perfusion time*

| **Primary study report** | **Allocation region** | **Subgroups** | **Donor type** | **Donor** | **Donor Age** | **Donor warm ischemia time** | **Perfusion time** |
| --- | --- | --- | --- | --- | --- | --- | --- |
| Matos 2017 ^6^ | Brazil | HMP | DBD | 54 | *42.5*  *(27.25-50.75)* | - | *11 (9.25-13.75)* |
| Moser 2017^7^ | Canada | HMP | DCD | 16 | 44.3 ± 14.1 | 46 ± 18 | NG |
| Sandal 2018^8^ | Canada | RR<0.4 | DBD | 151 | 55.86 ±12.8 | - | NG |
|  |  | RR<0.4 | DCD | 22 | 55.86 ± 12.8 | NG | NG |
|  |  | RR>0.4 | DBD | 15 | 59.93 ± 10.23 | - | NG |
|  |  | RR<0.2 | DBD | 113 | 54.63 ± 12.96 | - | NG |
|  |  | RR<0.2 | DCD | 12 | 54.63 ± 12.96 | NG | NG |
|  |  | RR>0.2 | DBD | 96 | 57.3 ± 12.37 | - | NG |
|  |  | RR>0.2 | DCD | 10 | 57.3 ± 12.37 | NG | NG |
| Zhong 2017^9^ | China | HMP | DCD | 141 | 32.8 ± 12.4 | *11.2 (5-25)* | *10.1 (4.1-16.0)* |
| Chen 2018^10^ | China | High RR | DCD | 22 | 54.7 ± 3.4 | 13.8 ± NG | NG |
|  |  | Low RR | DCD | 36 | 52.1 ± 5.3 | 11.1 ± NG | NG |
| Tai 2018^11^ | China | ECD^a^ | DCD | 446^b^ | 42.6 ± 14.4 | NG | NG |
|  |  | No ECD^a^ | DCD | 446^b^ | 42.6 ± 14.4 | NG | NG |
| Zheng 2020^13^ | China | NA | DCD | 181 | 50.8 ± 12.5 | 5.1 ± 2.2 | NG |
| Zeng 2021^15^ | China | RP | DBD | 15 | 50.67 ± 10.9 | - | 6.64 ± 3.87 |
|  |  | AP | DBD | 15 | 50.67 ± 10.9 | - | 5.25 ± 3.95 |
| Kievit 1998^16^ | Eurotransplant | NA | DCD | 66 | 44 (14-70) | 44 (9-80) | 8 ± NG |
| De Vries 2010^17^ | Eurotransplant | NA | DCD | 439 | 45 ± 16 | 26 ±11 | NG |
| Jochmans 2011^19^ | Eurotransplant | NA | DBD | 294 | 51 (16-81) | - | NG |
|  |  | NA | DCD | 42 | 51 (16-81) | NG | NG |
| Gallinat 2017^20^ | Eurotransplant | NA | DBD | 43 | *66 (28-82)* | - | *5.5 (1.6-12.7)* |
| Houtzager 2021^21^ | Eurotransplant | Donor 1 | DBD | 1 | NG | - | 15 |
|  |  | Donor 2 | DBD | 1 | NG | - | 6 |
|  |  | Donor 3 | DBD | 1 | NG | - | 15 |
|  |  | Donor 4 | DCD | 1 | NG | NG | 4 |
|  |  | Donor 5 | DBD | 1 | NG | - | 5 |
| Weberskirch 2022^22^ | Eurotransplant | NA | DBD | 45 | *56 (19-84)* | *-* | *10.1 (3.4-18.3)* |
|  |  |  | DCD | NG | *56 (19-84)* | *34 (16-65)* | *10.1 (3.4-18.3)* |
| Matillon 2017^23^ | France | NRP | DCD | 17 | 43.4 ± 9.8 | 128.1 ±15.3 | NG |
|  |  | In situ cold perfusion | DCD | 9 | 42.2 ± 12.2 | 135.9 ±17.5 | NG |
| Mori 2019^24^ | Italy | NA | DCD | 6 | 57.3 ± NG | NG | 4.47 ± 1.46 |
| Bissolati 2021^25^ | Italy | SKT | DBD | 31 | 68.3 ± 8.4 | - | 6.05 ± 3.06 |
|  |  | DKT | DBD | 26 | 76.9 ± 5.9 | - | 6.05 ± 3.06 |
|  |  | SCORE 0-4 | DBD | 42 | NG | - | 6.05 ± 3.06 |
|  |  | SCORE 5-7 | DBD | 41 | NG | - | 6.05 ± 3.06 |
| Ruberto 2022^28^ | Italy | NA | DBD | 23 | *67* | - | *4.11 (2.0-5.5)* |
| Offerni 2024^29^ | Italy | KDPI <50% | DCD | 34 | 49.38 ± 13.86 | 24.7 ±11.64 | NG |
|  |  | KDPI>50% | DCD | 38 | 60.26 ± 8.67 | 24.71 ±10.26 | NG |
| Matsuno 2006^30^ | Japan | flow 0.45-0.65 ml/min/g | DBD | 2 | 50.1 ± 12.6 | - | NG |
|  |  | flow 0.45-0.65 ml/min/g | DCD | 33 | 50.1 ± 12.6 | 19.3 ±31.4 | NG |
|  |  | flow 0.65-0.90 ml/min/g | DBD | 7 | 45.2 ± 12.3 | - | NG |
|  |  | flow 0.65-0.90 ml/min/g | DCD | 23 | 45.2 ± 12.3 | 8.2 ±12.1 | NG |
|  |  | flow > 0.9 ml/min/g | DBD | 5 | 41.6 ± 15.7 | - | NG |
|  |  | flow > 0.9 ml/min/g | DCD | 18 | 41.6 ± 15.7 | 12 ±30.7 | NG |
| Matsuno 2010^35^ | Japan | Age donor ≤ 50y | DCD | 9 | 34.1 ± 8.2 | 7 ±5.7 | 5.91 ± 4.9 |
|  |  | Age donor ≥ 50y | DCD | 8 | 54.9 ± 3.3 | 8.8 ±5.8 | 11.71 ± 6.8 |
| Iwamoto 2023^36^ | Japan | NA | DBD | 10 | 53.8 ± NG | - | 2.3 (1-4) |
|  |  | NA | DCD | 3 | 53.8 ± NG | NG | 2.3 (1-4) |
| Kwiatkowski 2001^37^ | Poland | IF | DBD | 143 | 36 ± 13.4 | - | NG |
|  |  | DGF | DBD | 84 | 36 ± 13.4 | - | NG |
|  |  | PNF | DBD | 7 | 36 ± 13.4 | - | NG |
| Wszola 2019^40^ | Poland | R1 RR<0.19 mm Hg/mL/min | DBD | 187 | 43.1 ± 14 | - | NG |
|  |  | R2 RR=>0.19 mm Hg/mL/min | DBD | 220 | 49.2 ± 16.4 | - | NG |
|  |  | R2-Ind+ | DBD | 124 | 49.2 ± 16.4 | - | NG |
|  |  | R2-Ind- | DBD | 96 | 49.2 ± 16.4 | - | NG |
| Elec 2014^42^ | Romania | NA | DBD | 82 | NG | - | NG |
| Suarez 1999^43^ | Spain | Donor 1 | DBD | 1 | 31 | - | 7 |
|  |  | Donor 2 | DBD | 1 | 73 | - | 6 |
|  |  | Donor 3 | DBD | 1 | 60 | - | 15 |
|  |  | Donor 4 | DBD | 1 | 54 | - | 17 |
|  |  | Donor 5 | DBD | 1 | 72 | - | 20 |
| Gomez 2015^44^ | Spain | NA | DBD | 93 | 73.6 ± NG | - | NG |
| Metcalfe 2001^46^ | United Kingdom | fit for transplantation | DCD | 8 | NG | NG | 6 ± NG |
|  |  | unfit for transplantation | DCD | 6 | NG | NG | 6 ± NG |
| Bellini 2019^47^ | United Kingdom | NA | DBD | 21 | 58 ± 14 | - | 5.7 ± 3.9 |
|  |  | NA | DCD | 12 | 58 ± 14 | NG | 5.7 ± 3.9 |
| Sevinc 2019^48^ | United Kingdom | pulsatile perfusion | DCD | 64 | 51 ± NG | 60.43 ±11.09 | NG |
|  |  | continuous perfusion | DCD | 64 | 51 ± NG | 60.56 ±16.23 | NG |
| Henry 1988^50^ | United States of America | primary machine perfused  + Belzer | NG | 140 | NG | NG | NG |
|  |  | secondary machine perfused + Belzer | NG | 14 | NG | NG | NG |
|  |  | machine perfused + silica gel | NG | 100 | NG | NG | NG |
| Tesi 1993^51^ | United States of America | age donor ≥ 60y | DBD | 50 | 64 ± NG | - | 14.6 local - 14.2 import |
|  |  | age donor ≥40 <60y | DBD | 37 | 48 ± NG | - | 14.6 local - 14.2 import |
| Polyak 2000a^53^ | United States of America | no ECD | DBD | 237 | 40.8 ± 8.8 | - | 12 ± NG |
|  |  | ECD | DBD | 165 | 54.8 ± 9.6 | - | 12 ± NG |
| Sonnenday 2003^57^ | United States of America | NA | DBD | 14 | 46 ±NG | - | 11 (6.8-20.1) |
| Mozes 2005^58^ | United States of America | RR <0.4 | NG | 221 | NG | NG | NG |
|  |  | RR 0.41-0.5 | NG | 44 | NG | NG | NG |
|  |  | RR 0.51-0.6 | NG | 15 | NG | NG | NG |
| Nyberg 2005^59^ | United States of America | NA | DBD | 425 | NG | - | NG |
| Cho 2008^60^ | United States of America | NA | NG | 678 | NG | NG | NG |
| Sung 2008^61^ | United States of America | NA | DBD | 7456 | NG | - | NG |
|  |  | NA | DCD | 1430 | NG | NG | NG |
| Nghiem 2009^62^ | United States of America | NA | DBD | 14 | 33.3 ± NG | - | 16.7 (7-24) |
| Hugen 2011^63^ | United States of America | Used | NG | 120 | 62 ± 10 | NG | NG |
|  |  | Discarded | NG | 188 | 57 ± 13 | NG | NG |
| Patel 2012a^65^ | United States of America | NH | DBD | 45 | 51 ± 14 | - | 12 ± 6.2 |
|  |  | NH | DCD | 12 | 51 ± 14 | NG | 12 ± 6.2 |
|  |  | AH | DBD | 55 | 57.6 ± 10 | - | 10.8 ± 5.4 |
|  |  | AH | DCD | 5 | 57.6 ± 10 | NG | 10.8 ± 5.4 |
| Yushkov 2012^66^ | United States of America | NA | DBD | 408 | 48.6 ± 12.5 | - | NG |
|  |  | NA | DCD | 46 | 48.6 ± 12.5 | NG | NG |
| Woodside 2015^67^ | United States of America | tPA | DCD | 12 | 49 ± NG | NG | NG |
|  |  | Control | DCD | 12 | 50.2 ± NG | NG | NG |
| Parikh 2016^68^ | United States of America | DGF | DBD | 146 | 46.7 ± 13 | - | *11 (7-15)* |
|  |  | DGF | DCD | 84 | 46.7 ± 13 | NG | *11 (7-15)* |
|  |  | No DGF | DBD | 350 | 46.9 ± 14.1 | - | *10 (6-14)* |
|  |  | No DGF | DCD | 91 | 46.9 ± 14.1 | NG | *10 (6-14)* |
| Mendez 2021^71^ | United States of America | DGF | NG | 79 | *49 (36-56)* | *27 (23-34)* | *13.1 (8.2-18.9)* |
|  |  | No-DGF | NG | 215 | *42 (30-55)* | *29 (24-36)* | *13.4 (8.4-19.7)* |
| Zhu 2021^72^ | United States of America | ENO | DBD | 7 | 51 (27-73) | - | 12 ± 9 |
|  |  | Control | DBD | 20 | 49 (35-70) | - | 8 ± 5 |
| Chotai 2023^73^ | United States of America | MP | DBD | 28 | *47* | - | *7.33 (4.5-10.38)* |
|  |  | MP | DCD | 3 | *47* | NG | *7.33 (4.5-10.38)* |
|  |  | GP | DBD | 1156 | *37* | - | *8.87 (5.65-12.25)* |
|  |  | GP | DCD | 125 | *37* | NG | *8.87 (5.65-12.25)* |
| Duarte 2023^74^ | United States of America | NA | DBD | 37 | 39 ± 15 | - | 7.95 ± 4.2 |
|  |  | NA | DCD | 12 | 39 ± 15 | 10.6 ±5.15 | 7.95 ± 4.2 |
| Werenski 2023^75^ | United States of America | MP | DBD | 60 | 47 ± 17 | - | 13 ± 7 |
|  |  | MP | DCD | 31 | 47 ± 17 | 25 ±9 | 13 ± 7 |
|  |  | GP | DBD | 416 | 44 ± 15 | - | 12 ± 6 |
|  |  | GP | DCD | 182 | 44 ± 15 | 24 ±11 | 12 ± 6 |

^a^ In China, DCD is the only source of the deceased donor at present. ^b^ Only the whole population is given. Median values are given in italics.

AH, abnormal histology; AP, anterograde perfusion; DGF, kidney delayed graft function; DKT, double kidney transplantation; ECD, expanded criteria donor; ENO, S-ethyl nitrite; IF, immediate function; KDPI, Kidney Donor Profile Index; MP, marginal perfusion; NG, not given; NH, normal histology; PNF, primary non function; RP, retrograde perfusion; RR, renal resistance; SKT, single kidney transplantation; tPA, tissue plasminogen activator; NRP, normothermic regional perfusion; GP, good perfusion.

*Table S3 Overview of primary study reports that preselected kidneys based on perfusion parameters introducing a selection bias.*

| **Primary study report** | **Preselection perfusion parameter cut-off** | **Time Point** |
| --- | --- | --- |
| Mendez 2021^71^ | RR >0.3mmHg/mL/min | Terminal |
| Matillon 2017^23^ | RR >0.5mmHg/ml/min | 6h |
| Patel 2012^65^ | Flow <59.7 mL/min and Initial RR >0.57 mmHg/ml/min | Initial |
| Nghiem 2009^62^ | Flow <47 mL/min and RR > 0.4mmHg/ml/min | Terminal |
| Matsuno 2006^30^ | Flow <0.4ml/min/g with a rising perfusion pressure pattern | Not specified |
| Nyberg 2005^59^ | RI >0.55 RI was calculated by the quotient of mean perfusion pressure (mmHg) divided by perfusate flow rate (ml/min/100 g renal mass). | Terminal |
| Tesi 1993^51^ | RR>=0.4mmHg/ml/min or flow <70 ml/min were discarded | Not specified |

*Table S4 Perfusion devices*

| Device | Pump type | Pulsatile | Pump control |
| --- | --- | --- | --- |
| LifePort | Roller pump | Yes | Pressure |
| Airdrive HMP-system | Centrifugal pump | Yes | Pressure |
| CMP-X08 | Centrifugal pump | No | Pressure |
| Gambro PF-3B | Roller pump | Yes | Flow |
| Kidney Assist Transport | Centrifugal pump | Yes | Pressure |
| RM3 | Roller pump | Yes | Pressure |
| LPS-II | Centrifugal pump | No | NG |
| MOX100 | Centrifugal pump | Yes | Pressure |
| Waves | Centrifugal pump | Yes | Flow |

*Table S5 Pump control adaptation*

| **Primary study report** | **Pump control adapted** | **Changed Value** | | **Adaptation** | **Oxygen** | **Additives** |
| --- | --- | --- | --- | --- | --- | --- |
|  |  | **Flow** | **Pressure** |  |  |  |
| Matos 2017^6^ | NG | NA | NA | NA | No | NG |
| Moser 2017 ^7^ | NG | NA | NA | NA | No | NG |
| Sandal 2018 ^8^ | Yes | No | Yes | Reduced to 25 mmHg if flow rates were extremely high | No | Mannitol 2.5g/l |
| Zhong 2017^9^ | No | NA | NA | NA | No | NG |
| Chen 2018^10^ | NG | NA | NA | NA | NG | NG |
| Tai 2018^11^ | Yes | No | Yes | After 1 to 1.5 hour, if the flow was <80 mL/min, it was gradually increased (upper limit, 45 mm Hg) to increase the flow to 80-120 mL/min, and if the flow was >140 mL/min, the pressure was decreased to keep the flow at 100-140 mL/min. | No | NG |
| Zheng 2020^13^ | Yes | No | Yes | After a half-hour, if the flow was 140 mL/min, pressure was decreased to maintain 100–140 mL/min | No | NG |
| Zeng 2021^15^ | Yes | No | Yes | The initial perfusion pressure was set at 15 mm Hg, if went well perfusion pressure was gradually reduced to 12 mm. 1mmHg lower every 10min otherwise was maintained at 15 mmHg | No | NG |
| Kievit 1998^16^ | NG | NA | NA | NA | NG | NG |
| De Vries 2010^17^ | Yes | No | Yes | During the first hour to keep pressure at 55 mmHg | NG | 40U Insulin; 200 000U penicillin; 16mg dexamethasone; sodium bicarbonate if needed |
| Jochmans 2011^19^ | No | NA | NA | NA | No | NG |
| Gallinat 2017^20^ | NG | NA | NA | NA | No | NG |
| Houtzager 2021^21^ | No | NA | NA | NA | Yes | NG |
| Weberskirch 2022^22^ | NG | NA | NA | NA | No | NG |
| Matillon 2017^23^ | NG | NA | NA | NA | No | NG |
| Mori 2019^24^ | NG | NA | NA | NA | Yes | NG |
| Bissolati 2021^25^ | NG | NA | NA | NA | Yes | NG |
| Ruberto 2022^28^ | NG | NA | NA | NA | No | NG |
| Offerni 2024^29^ | NG | NA | NA | NA | No | NG |
| Matsuno 2006^30^ | NG | NA | NA | NA | Yes | NG |
| Matsuno 2010^35^ | NG | NA | NA | NA | Yes | NG |
| Iwamoto 2023^36^ | NG | NA | NA | NA | Yes (in 4 cases) | No |
| Kwiatkowski 2001^37^ | NG | NA | NA | NA | NG | NG |
| Wszola 2019^40^ | NG | NA | NA | NA | No | NG |
| Elec 2014^42^ | NG | NA | NA | NA | No | NG |
| Suarez 1999^43^ | NG | NA | NA | NA | No | 40 UI insulin, 8mg dexamethasone, 4mg Trifluoperazine |
| Gomez 2015^44^ | NG | NA | NA | NA | No | NG |
| Metcalfe 2001^46^ | NG | NA | NA | NA | NG | NG |
| Bellini 2019^47^ | Yes | No | Yes | Initial peak systolic pressure of 45mmHg. After 30 min cold perfusion pressure was held constant at 40 mmHg. | No | NG |
| Sevinc 2019^48^ | No | NA | NA | NA | No | NG |
| Henry 1988^50^ | NG | NA | NA | NA | NG | NG |
| Tesi 1993^51^ | NG | NA | NA | NA | NG | Mannitol; sodium bicarbonate, CO2; verapamil (rarely priscoline) |
| Polyak 2000a^53^ | NG | NA | NA | NA | NG | Prostaglandin, trifluoperazine, verapamil, papaverine vs control |
| Sonnenday 2003^57^ | NG | NA | NA | NA | NG | Mannitol, sodium, bicarbonate, CO2, verapamil |
| Mozes 2005^58^ | Yes | No | Yes | Pump pressures were set at 60 mm Hg and allowed to decrease | NG | NG |
| Nyberg 2005^59^ | NG | NA | NA | NA | NG | Mannitol, sodium bicarbonate, CO2, verapamil |
| Cho 2008^60^ | NG | NA | NA | NA | NG | NG |
| Sung 2008^61^ | NG | NA | NA | NA | NG | NG |
| Nghiem 2009^62^ | NG | NA | NA | NA | NG | tPA 200mg |
| Hugen 2011^63^ | NG | NA | NA | NA | NG | NG |
| Patel 2012a^65^ | NG | NA | NA | NA | No | NG |
| Yushkov 2012^66^ | NG | NA | NA | NA | No | NG |
| Woodside 2015^67^ | Yes | No | Yes | If flow rates were <100 mL/min, pressure  was increased by 5 mmHg and 10 mg of verapamil  hydrochloride. If flows remained <100 mL/min,  pressure was increased by 5 mmHg one final time | No | 50 mg Alteplase 40 U insulin 200 000 U penicillin G 16 mg  dexamethasone |
| Parikh 2016^68^ | NG | NA | NA | NA | No | No |
| Mendez 2021^71^ | NG | NA | NA | NA | No | NG |
| Zhu 2021^72^ | NG | NA | NA | NA | No | No |
| Chotai 2023^73^ | NG | NA | NA | NA | No | NG |
| Duarte 2023^74^ | NG | NA | NA | NA | No | NG |
| Werenski 2023^75^ | Yes | No | Yes | Target flow >100 mL/min and resistance <0.30 mmHg/mL/min was maintain with Mannitol (1 g) and vasodilators verapamil (10 mg, max 3 doses), papaverine (10 mg, max 3 doses), phentolamine (5 mg, max 3 doses) | No | Penicillin (250,000 U), insulin (40 U), dexamethasone  (16 mg), and mannitol (3 g). |

*Table S6 Perfusion characteristics*

| **Primary study report** | **Device** | **Temperature (°C)** | **Pump** | **Pulsatile** | **Pump control** | **Control adapted** | **Perfusion Solution** | **Additives** | **Perfusion strategy** |
| --- | --- | --- | --- | --- | --- | --- | --- | --- | --- |
| Matos 2017^6^ | LifePort | 1–8 | Roller pump | Yes | Pressure | NG | UW Machine perfusion solution | NG | Back to base |
| Moser 2017^7^ | LifePort | 2-4 | Roller pump | Yes | Pressure | NG | UW Machine perfusion solution | NG | Continuous |
| Sandal 2018^8^ | LifePort | NG | Roller pump | Yes | Pressure | Yes | UW Machine perfusion solution | Yes | Back to base |
| Zhong 2017^9^ | LifePort | 0-4 | Roller pump | Yes | Pressure | No | UW Machine perfusion solution | NG | Continuous |
| Chen 2018^10^ | LifePort | NG | Roller pump | Yes | Pressure | NG | NG | NG | Continuous |
| Tai 2018^11^ | LifePort | NG | Roller pump | Yes | Pressure | Yes | NG | NG | Continuous |
| Zheng 2020^13^ | LifePort | NG | Roller pump | Yes | Pressure | Yes | NG | NG | Continuous |
| Zeng 2021^15^ | LifePort | 1-8 | Roller pump | Yes | Pressure | Yes | NG | NG | NG |
| Kievit 1998^16^ | Gambro PF-3B | 4 | Roller pump | Yes | Flow | NG | UW Machine perfusion solution | NG | Continuous |
| De Vries 2010^17^ | Gambro PF-3B | 4 | Roller pump | Yes | Flow | Yes | UW Gluconate | Yes | Continuous |
| Jochmans 2011^19^ | LifePort | 1–8 | Roller pump | Yes | Pressure | No | UW Machine perfusion solution | NG | Continuous |
| Gallinat 2017^20^ | LifePort | 1-8 | Roller pump | Yes | Pressure | NG | HTK OR UW Machine perfusion solution | NG | Back to base |
| Houtzager 2021^21^ | Airdrive HMP System | 8 -12 | Centrifugal pump | Yes | Pressure | No | UW Machine perfusion solution | NG | Back to base |
| Weberskirch 2022^22^ | LifePort | 2-4 | Roller pump | Yes | Pressure | NG | UW Machine perfusion solution | NG | NG |
| Matillon 2017^23^ | RM3 | 1–4 | Roller pump | Yes | Pressure | NG | UW Machine perfusion solution | NG | Continuous |
| Mori 2019^24^ | Kidney Assist Transport | NG | Centrifugal pump | Yes | Pressure | NG | NG | NG | Continuous |
| Bissolati 2021^25^ | Waves | 5+-2 | Centrifugal pump pump | Yes | Flow | NG | UW Machine perfusion solution | NG | Continuous |
| Ruberto 2022^28^ | LifePort | NG | Roller pump | Yes | Pressure | NG | UW Machine perfusion solution | NG | Continuous/Back to base |
| Offerni 2024^29^ | LifePort | 3-7 | Roller pump | Yes | Pressure | NG | UW Machine perfusion solution | NG | Continuous |
| Matsuno 2006^30^ | LPS-II, Nikkiso, Tokyo, Japan | 8-10 | Centrifugal pump | No | NG | NG | cryoprecipitated AB-positive human plasma | NG | Back to base |
| Matsuno 2010^35^ | LPS-II, Nikkiso, Tokyo, Japan | 8-10 | Centrifugal pump | No | NG | NG | cryoprecipitated AB-positive human plasma or UW-Gluconate | NG | Continuous |
| Iwamoto 2023^36^ | CMP-X08 | 4-8 | Centrifugal pump | No | Pressure | NG | UW machine perfusion solution | No | NG |
| Kwiatkowski 2001^37^ | MOX100 | 6 | Centrifugal pump | Yes | NG | NG | UW Gluconate | NG | Back to base |
| Wszola 2019^40^ | LifePort | 4-6 | Roller pump | Yes | Pressure | NG | UW Machine perfusion solution | NG | Back to base |
| Elec 2014^42^ | LifePort | 2-6 | Roller pump | Yes | Pressure | NG | UW Machine perfusion solution | NG | NG |
| Suarez 1999^43^ | MOX100 | 5-7 | Centrifugal pump | Yes |  | NG | UW Machine perfusion solution | Yes | Back to base |
| Gomez 2015^44^ | LifePort | NG | Roller pump | Yes | Pressure | NG | UW Machine perfusion solution | NG |  |
| Metcalfe 2001^46^ | NG | 3-8 | NG | NG | NG | NG | hyperosmolar citrate solution | NG | Continuous |
| Bellini 2019^47^ | RM3 | 4-5 | Roller pump | Yes | Pressure | Yes | UW Machine perfusion solution | NG | Back to base |
| Sevinc 2019^48^ | LifePort | NG | Roller pump | Yes | Pressure | No | UW Machine perfusion solution | NG | NG |
| Henry 1988^50^ | MOX100 | NG | Centrifugal pump | Yes | NG | NG | UW Machine perfusion solution | NG | Back to base |
| Tesi 1993^51^ | MOX100 | NG | Centrifugal pump | Yes | NG | NG | UW Machine perfusion solution | Yes | Continuous/Back to base |
| Polyak 2000a^53^ | MOX100 / RM3 | 4 | Centrifugal pump | Yes | Pressure | NG | UW Machine perfusion solution | Yes | Continuous |
| Sonnenday 2003^57^ | LifePort | 5 | Roller pump | Yes | NG | NG | UW Machine perfusion solution | Yes | Back to base |
| Mozes 2005^58^ | RM3 | NG | Roller pump | NG | NG | Yes | UW Machine perfusion solution | NG | Continuous |
| Nyberg 2005^59^ | MOX100 | NG | Centrifugal pump | Yes | Pressure | NG | NG | Yes | Continuous |
| Cho 2008^60^ | NG | NG | NG | Yes | NG | NG | NG | NG | NG |
| Sung 2008^61^ | NG | NG | NG | NG | NG | NG | NG | NG | NG |
| Nghiem 2009^62^ | RM3 | 4.2 (3.2-5.1) | Roller pump | Yes | NG | NG | HTK | Yes | Continuous |
| Hugen 2011^63^ | LifePort / RM3 | NG | Roller pump | Yes | Pressure | NG | NG | NG | NG |
| Patel 2012a^65^ | LifePort | NG | Roller pump | Yes | Pressure | NG | UW Machine perfusion solution | NG | Continuous |
| Yushkov 2012^66^ | LifePort | NG | Roller pump | Yes | Pressure | NG | NG | NG | Continuous/Back to base |
| Woodside 2015^67^ | RM3 | 4–6 | Roller pump | Yes | Pressure | Yes | IGL Pulsatile Perfusion Solution | Yes | Continuous |
| Parikh 2016^68^ | LifePort | 4 | Roller pump | Yes | Pressure | NG | UW Machine perfusion solution | No | Continuous |
| Mendez 2021^71^ | RM3 | 4 | Roller pump | Yes | Pressure | NG | UW Machine perfusion solution | NG | Back to base |
| Zhu 2021^72^ | LifePort | 4 | Roller pump | Yes | Pressure | NG | NG | Yes | NG |
| Chotai 2023^73^ | LifePort | 4.90 (4.75-5.20) | Roller pump | Yes | Pressure | NG | NG | NG | Continuous |
| Duarte 2023^74^ | LifePort | NG | Roller pump | Yes | Pressure | NG | UW Machine perfusion solution | NG | NG |
| Werenski 2023^75^ | LifePort | 5 | Roller pump | Yes | Pressure | Yes | UW Machine perfusion solution | Yes | NG |

HTK, histidine-tryptophane-ketoglutarate; IGL, Institut Georges Lopez; NG, not given; UW, University of Wisconsin

Table S7 Histological findings and their association with renal resistance (RR) during hypothermic machine perfusion

| Study | Histology Feature | Timepoint | Findings | Outcome Association |
| --- | --- | --- | --- | --- |
| Bissolati 2021^25^ | Karpinski score | Baseline | Baseline RR was 2.71 (score 0-4) vs. 3.35 (score 5-7) (p=0.074) | No direct link to post-transplant outcomes |
| Bissolati 2021^25^ | Karpinski score | 1h | At 60 min: RR 0.94 (score 0-4) vs. 1.24 (score 5-7) (p=0.031) | ROC-derived RR threshold: ≥0.88 at 120 min (sensitivity 0.71, specificity 0.75) |
| Bissolati 2021^25^ | Karpinski score | 2h | At 120 min: RR 0.81 (score 0-4) vs. 1.10 (score 5-7) (p=0.010) |  |
| Zheng 2020^13^ | Remuzzi score | Terminal | Weak, but significant correlation between RR and histology (RI mmHg/(mL/min)= 0.356, p < 0.001) | Correlation with DGF occurrence |
| Zheng 2020^13^ | ATI | Terminal | No correlation between ATI and RR (RI mmHg/(mL/min) = 0.053, p = 0.335) but ATI strongly predicted DGF (OR 4.72, p < 0.001) | DGF occurrence post-transplant |
| Patel 2012^64^ | Glomerulosclerosis and hyaline arteriosclerosis | Mean & terminal | Higher RR was observed in kidneys with glomerulosclerosis and hyaline arteriosclerosis, but correlation strength was weak. | Higher RR associated with worse histology but no outcome data |
| Yushkov 2012^66^ | TIS | 1.5h, 3h, 5h | TIS at 26%-50% significantly increased odds of RR >0.3 mmHg/mL/min (p=0.01) | TIS was the most predictive histological factor for high RR and worse renal outcomes |
| Yushkov 2012^66^ | Arterial IFN & glomerulosclerosis | 1.5h, 3h, 5h | IFN and glomerulosclerosis not increase odds of higher RR | No significant relationship between IFN, glomerulosclerosis and changes in RR |

ATI, acute tubular injury; DGF, delayed graft function; RR, renal resistance; ROC, receiver operating characteristic; TIS, tubular interstitial scarring; IFN, intimal fibrous narrowing.

Table S8 Renal resistance threshold studies: summary of findings and robustness grading

| Study | Perfusion device | Findings | Robustness level |
| --- | --- | --- | --- |
| Kozaki 2000^34^ | LPS-III | Average RR > 80 mmHg/mL/min/g was used as a discard threshold (based on prior experimental work). Kidneys above this threshold were discarded. Contralateral kidneys, transplanted at other centers, developed PNF — but their perfusion parameters were not reported. No ROC analysis, multivariable modeling, or validation was performed. | Low |
| Nyberg 2005^59^ | MOX100 | Terminal RR > 0.4 mmHg/mL/min was associated with higher serum creatinine at 1 year (ANOVA), but was not statistically associated with DGF or graft survival (no ROC or regression analysis). | Low |
| Yushkov 2012^66^ | LifePort | RR> 0.3 mmHg/mL/min at 3 and 5h is associated with lower 1-year graft survival (Kaplan-Meier, log-rank test; Cox regression). | High |
| Gomez 2015^44^ | LifePort | Terminal RR ≥ 0.3 mmHg/mL/min was evaluated as a threshold for DGF prediction (ROC AUC = 0.58), but RR was not significantly associated with DGF in multivariable analysis. | Low |
| Burgos Revilla  2015^45^ | LifePort | Terminal RR ≥ 0.3 mmHg/mL/min was investigated as a threshold for DGF prediction (ROC analysis: AUC 0.58 (95% CI: 0.44–0.71), sensitivity 74%, specificity 53%, positive predictive value = 29%, negative predictive value 89%); however, RR was not significantly associated with DGF or graft survival (multivariable Cox regression). | Low |
| Sandal 2018^8^ | LifePort | Terminal RR ≥ 0.2 and ≥ 0.4 mmHg/mL/min were associated with increased death-censored graft failure (Cox proportional hazards regression; Kaplan-Meier; ROC-informed thresholds). DGF was not significantly associated with terminal RR. | High |
| Bissolati 2018^26^ | Waves | Final RR > 1.0 mmHg/mL/min at 3 hours was used as a discard threshold, and RR ≤ 1.0 within 60 minutes was associated with reduced PNF/DGF and faster creatinine decline (t-tests; chi-square; no ROC or regression analysis). | Low |
| Chen 2018^10^ | LifePort | RR > 0.4 mmHg/mL/min at 1h is associated with increased DGF and worse 1-year renal function (ROC analysis: sensitivity 62%, specificity 82%, Chi² test; t-test). | Low |
| Bellini 2019^47^ | RM3 | RR ≥ 0.45 mmHg/mL/min at 2h predicted DGF in DCD kidneys (ROC analysis: AUC 0.78, sensitivity 75%, specificity 80%); RR ≥ 0.2 mmHg/mL/min at 2h predicted DGF in DBD kidneys (ROC analysis: AUC 0.87, sensitivity 100%, specificity 91%). | Low |
| Bissolati 2019^27^ | Waves | RR ≤ 1.0 mmHg/mL/min within 3h was as an outcome predictor (time to reach threshold) (Chi² and t-tests applied) but RR was not statistically associated with DGF or long-term graft outcomes (Mann-Whitney U test; descriptive analysis). | Low |
| Wszola 2019^40^ | LifePort | RR ≥ 0.19 mmHg/mL/min at 4h is associated with increased acute rejection and lower 1-year graft survival (ROC analysis; logistic regression). | Moderate |
| Qiao 2021^14^ | LifePort | Terminal RR ≥0.30 mmHg/mL/min is associated with increased DGF (multivariable logistic regression; ROC analysis: 0.76 (95% CI 0.69-0.82) based on Youden index), but not with long-term graft survival (not evaluated); it is also associated with delayed renal function recovery (Cox regression). | Moderate |
| Mendez 2021^71^ | RM3 | Terminal RR ≥ 0.23 mmHg/mL/min is associated with increased DGF (logistic regression; ROC analysis Youden Index but no predictive metrics). | Low |
| Ruberto 2022^28^ | LifePort | Initial RR ≥0.5 mmHg/mL/min predicts DGF (ROC analysis AUC 0.083; sensitivity 82%, specificity 83%; Fisher’s exact test) but not long-term graft survival (Kaplan-Meier and log-rank test). | Low |
| Chotai 2023^73^ | LifePort | Terminal RR>0.4 is associated with increased DGF (Chi² test) and reduced long-term graft survival (Kaplan-Meier with log-rank). | Moderate |
| Werenski 2023^75^ | LifePort | Terminal RR ≥ 0.40 mmHg/mL/min independently predicts inferior long-term graft survival (multivariable Cox-regression). | Low |
| Offerni 2024^29^ | LifePort | Combination of average terminal low terminal RR (<0.15) and high flow (>150 mL/min) was associated with better 30d eGFR (ANOVA, Tukey post-hoc). No predictive modeling or threshold derivation performed. | Low |

AUC, area under the curve; CI, confidence interval; DCD, donation after circulatory death; DBD, donation after brain death; DGF, delayed graft function; eGFR, estimated glomerular filtration rate; HR, hazard ratio; PNF, primary non-function; PPV, positive predictive value; ROC, receiver operating characteristic; RR, renal resistance.

Table S9 Criteria-based grading of study robustness in RR threshold research

| Study | Robustness level | Threshold Derivation | Association Tested | Multivariable Adjustment | Predictive Performance | Sample Size | Selection Bias | Validation |
| --- | --- | --- | --- | --- | --- | --- | --- | --- |
| Kozaki 2000^34^ | Low | Arbitrary | No | No | No | Small | Present | No |
| Nyberg 2005^59^ | Low | No | Yes | No | No | Large | Present | No |
| Yushkov 2012^66^ | High | Yes | Yes | Yes | Yes | Large | No | Yes (internal) |
| Burgos Revilla  2015^45^ | Low | No | Yes | No | Limited | Likely adequate | No | No |
| Gomez 2015^44^ | Low | No | Yes | No | Limited | Adequate | No |  |
| Chen 2018^10^ | Low | No | No | No | No | Unclear | No |  |
| Sandal 2018^8^ | High | Yes | Yes | Yes | Yes | Large | No |  |
| Bissolati 2018^26^ | Low | Plausible | Yes | No | Limited | Small | No |  |
| Wszola 2019^40^ | Moderate | Yes | Yes | Limited | Yes | Large | No |  |
| Bellini 2019^47^ | Low | No | No | No | No | High | No |  |
| Bissolati 2019^27^ | Low | Plausible | Yes | No | Limited | Small | No |  |
| Qiao 2021^14^ | Moderate | Plausible | Yes | No | Yes | Adequate | No |  |
| Mendez 2021^71^ | Low | No | Yes | No | No | High | Present |  |
| Ruberto 2022^28^ | Low | Yes | Not reported | Yes | No | Unclear | No |  |
| Werenski 2023^75^ | Low | Not derived | Yes | No | No | Unclear | No |  |
| Chotai 2023^73^ | Moderate | Plausible (literature) | Yes | No | No | Likely adequate | No |  |
| Offerni 2024^29^ | Low | No | No | No | No | Unclear | No |  |

*Table S10 Essential data elements for transplant registries integrating machine perfusion*

| Parameter | Details | Explanation |
| --- | --- | --- |
| *Related to the donor* |  |  |
| Donor Characteristics | Age, sex, BMI, cause of death (DBD vs. DCD and warm ischemia time if DCD), comorbidities (hypertension, diabetes, cardiovascular disease), pre-retrieval creatinine, eGFR, urine output | Essential for contextualizing perfusion-related findings and risk stratification. |
| Kidney Weight | Measured pre-perfusion | May influence RR values and needs further study. |
| Biopsy Data | Histological scoring (fibrosis, arteriosclerosis, inflammation) of pre-implantation biopsy. | Provides insight into graft quality and transplantability. |
| *Related to the perfusion* | | |
| Perfusion Modality | Hypothermic (HMP) vs. Normothermic (NMP) | Different perfusion methods influence renal resistance and outcomes. |
| Device Used | LifePort, OrganOx, Wave, etc. | Device-dependent variability in RR values requires standardization. |
| Perfusion mode | Continuous, back-to-base | May influence RR trajectory and outcome. |
| Perfusion Duration | Total time on MP before transplantation | Prolonged MP duration may influence post-transplant outcomes. |
| Perfusate Composition | Type of perfusate (e.g. UW-MPS) and additives | Additives can alter perfusion dynamics and renal resistance. |
| Perfusion Temperature | Recorded continuously if available | Affects vascular compliance and resistance. |
| Oxygenated | Yes/No | May influence outcome. |
| Pulsatile vs. Continuous Flow | Type of perfusion mode used | Pulsatile flow may influence variability in RR. |
| Perfusion Pressure | Arterial pressure settings (e.g., 25 mmHg vs. 30 mmHg) | Necessary to standardize RR comparisons. |
| Flow and Pressure Readings | Arterial flow (ml/min), perfusion pressure (mmHg) to calculate RR. | Required for accurate RR calculation and comparison. |
| Essential Time Points | At the start of perfusion, at 15 minutes, 1 hour, and terminal measurement. Ideally all time points. | These time points capture the exponential decay trajectory of RR, ensuring key phases of stabilization are documented. |
| Cold Ischemia Time (CIT) | Time from cross-clamp to MP start | Delayed MP initiation may impact outcomes. |
| *Related to Recipient* |  |  |
| Recipient Age & Sex | Baseline demographic information | Helps assess risk stratification and outcome variability. |
| Recipient BMI | Body mass index at transplant | Obesity may impact post-transplant function and recovery. |
| Pre-Transplant Dialysis | Dialysis modality, duration, and last dialysis session | Prolonged dialysis before transplant may influence DGF risk. |
| Recipient Comorbidities | Hypertension, diabetes, cardiovascular disease | These factors influence graft function and long-term outcomes. |
| Immunological Matching | HLA mismatch level, panel reactive antibodies (PRA) | Important for rejection risk and long-term graft survival. |
| Immunosuppression Regimen | Type and dose of induction and maintenance therapy | Impacts graft survival and rejection rates. |
| *Outcome post-transplantation* | | |
| Primary Non-Function (PNF) | Yes/No outcome | Key post-transplant outcome measure. |
| Delayed Graft Function (DGF) | Yes/No, number of dialysis sessions required | Key post-transplant outcome measure. |
| eGFR Post-Transplant | Measured at 1, 3, and 12 months | Key post-transplant outcome measure. Helps assess long-term graft function. |
| Graft Loss & Patient Survival | Outcome data categorized by rejection vs. other causes | Key post-transplant outcome measure. |

BMI, body mass index; CIT, cold ischemia time; DBD, donation after brain death; DCD, donation after circulatory death; DGF, delayed graft function; eGFR, estimated glomerular filtration rate; HLA, human leukocyte antigen; MP, machine perfusion; PNF, primary non-function; PRA, panel reactive antibodies; RR, renal resistance.
